# Representativeness of participants in a randomized controlled trial on modalities of monitoring oral HIV pre-exposure prophylaxis use

**DOI:** 10.1101/2025.09.15.25335748

**Authors:** Marije L. Groot Bruinderink, Maarten F. Schim van der Loeff, Florien Dusseldorp, Katja van der Velde, Laura Blitz, Jean-Marie Brand, Colette C. A. J. van Bokhoven, Joey Woudstra, Koenraad Vermey, Sophie Boers, Maudy Sluimer, Hannelore M. Götz, Anders Boyd, Lotte Werner, Maaike L. Soors d’ancona, Frenk van Harreveld, Maria Prins, Elske Hoornenborg, Udi Davidovich, Vita W. Jongen

## Abstract

**Background:** We assessed whether participants in the EZI-PrEP study, a non-inferiority trial evaluating online and six-monthly monitoring of oral HIV pre-exposure prophylaxis (PrEP), represent the broader population of PrEP users in the Netherlands.

**Methods:** We conducted a cross-sectional study using routinely collected data from September 2021 to August 2022 at four sexual health centres (SHCs) in the Netherlands. Socio-demographic characteristics, sexual behaviour, and prevalence of bacterial STIs were compared between EZI-PrEP participants at baseline and other PrEP users during their first PrEP monitoring visit in said time period.

**Results:** The analysis included 469 EZI-PrEP participants and 5196 other PrEP users; respectively 99% and 96% were men who have sex with men (MSM). EZI-PrEP participants were less often transgender or gender divers persons (TGDP) (1% vs. 4%, p<0.001), older (median age=36 vs. 34 years, p=0.004), more often born in the Netherlands (68% vs. 58%, p<0.001), and more often completed a university/college degree (81% vs. 76%, p=0.01). They reported more group sex (38% vs. 33%, p=0.023) and condomless anal sex (95% vs. 92%, p=0.004), but less often sex work (1% vs. 6%, p<0.001). Prevalence of bacterial STIs was similar between groups (19% vs 18%, p=0.766).

**Conclusions:** The similar STI prevalence suggests comparable risk for HIV acquisition among EZI-PrEP participants and other PrEP users, making EZI-PrEP study outcomes applicable to a broader population of PrEP users. However, under-representation of TGDPs, sex workers, individuals not born in the Netherlands, and individuals without university or college degree may limit generalizability.

## 1. Introduction

Oral HIV pre-exposure prophylaxis (PrEP) is highly effective against HIV acquisition and is an important tool in curbing the HIV epidemic^1–5^. However, uptake and retention in PrEP programs is suboptimal^6–9^. Reported barriers to in-clinic PrEP care include anticipating or experiencing stigma, discrimination and lack of privacy at the PrEP providing facility, having difficulties with taking time off from work or other obligations to attend PrEP monitoring visits, travel costs^9–17^, and limited capacity of PrEP programmes due to staff shortages or budget limitations^18,19^. Some of these barriers could potentially be mitigated by reducing the frequency of in-clinic PrEP monitoring visits, for example, through online PrEP monitoring or less frequent in-clinic visits. While both approaches have been implemented in various formats and settings worldwide, their effect on PrEP adherence among PrEP users overall remains understudied.

The EZI-PrEP study was initiated to assess whether reducing the number of in-clinic PrEP monitoring visits would affect PrEP adherence. Designed as a 2×2 factorial, non-inferiority, randomized-controlled trial (RCT), the study aimed to determine, in terms of PrEP adherence, whether 6-monthly and online PrEP monitoring are non-inferior to 3-monthly and in-clinic PrEP monitoring^20^. The randomized nature of the EZI-PrEP study allows to control for differences in both measured and unmeasured characteristics between study arms and as such, enables to establish a causal relationship between the modalities of PrEP monitoring and adherence. Moreover, RCTs are known to commonly have selective eligibility criteria and stringent protocols, which are helpful in reducing statistical noise and ensuring appropriate assessment of both exposures and study outcomes (i.e., internal validity)^21–23^. However, applying selective criteria and stringent protocols might come at the cost of studying populations that are not representative of the broader target population (i.e., reducing generalizability)^21,22^. Assessments of potential issues in generalizability are rarely conducted for RCTs, since data on the target population are often lacking.

The EZI-PrEP study was nested within the much larger, government-funded National PrEP Programme (NPP), which offers free-of-charge PrEP care and subsidized PrEP tablets to 8,500 persons at increased risk for HIV acquisition [e.g., men who have sex with men (MSM) and transgender and gender diverse persons]^24^. Data on demographics, sexual health and sexually transmitted infections (STIs) are routinely collected as part of this programme. Assuming PrEP users of the NPP closely represent the target population of all PrEP users, comparing data between EZI-PrEP and the NPP could allow assessment of potential issues in generalizability of the EZI-PrEP trial.

The aim of the current study was therefore to compare the distributions of socio-demographic characteristics, sexual behaviour, and prevalence of bacterial STIs of EZI-PrEP study participants to other PrEP users, not enrolled in the study, who were seeking PrEP care at the same sites. Additionally, this study compares participant characteristics across the four study sites to identify potential site-specific differences.

## 2. Methods

### 2.1 Study setting

The EZI-PrEP study was embedded in the government-funded Dutch National PrEP pilot Programme (NPP)^24^. The NPP operated between 1 July 2019 to 31 July 2024 and was the most common route to receive PrEP care in the Netherlands. The NPP offered free-of-charge PrEP care and subsidized PrEP tablets to persons at increased risk for HIV aqcuisition^24^. The NPP was implemented by 23 sexual health centres of public health services in the Netherlands. The NPP was capped at 8,500 PrEP users; for each sexual health centre a maximum number of PrEP users was set. At the start of the EZI-PrEP study, waiting lists for NPP enrolment were in place. Four large sexual health centres (SHC), together accounting for approximately 40% of the NPP’s total capacity, participated in the EZI-PrEP study. These SHCs were located in the cities of Amsterdam, Rotterdam, The Hague and Nijmegen and served PrEP users in the respective public health service catchment areas.

### 2.2 Design, procedures and population

We conducted a cross-sectional analysis using routinely collected data on socio-demographics, sexual behaviour, PrEP use, and the testing and diagnosis of bacterial STIs. These data were derived from PrEP initiation and monitoring consultations following standardized procedures of the NPP. PrEP consultations include tailored counselling on PrEP use and sexual health, using motivational interviewing techniques. Each PrEP consultation also included screening for HIV and bacterial STIs (i.e., *Chlamydia trachomatis*, *Neisseria gonorrhoeae*, and *Treponema pallidum*). Testing for *Chlamydia trachomatis* and *Neisseria gonorrhoeae* was conducted at three anatomical sites (rectal, urogenital, and pharyngeal). Participants diagnosed with an STI were offered free-of-charge treatment in accordance with standard PrEP care protocols. A pre-defined selection of routinely collected data is pseudonymized and sent to the National STI surveillance database of the National Institute for Public Health and the Environment (RIVM, Bilthoven, the Netherlands) for monitoring and surveillance purposes. Individuals participating in the NPP may opt-out of sharing their data with this national STI surveillance database. After a PrEP consultation, individuals could obtain PrEP; they were given the number of tablets needed until their next consultation, based on their planned dosing schedule (i.e., daily or intermittent).

In this analysis we defined two groups of PrEP users: EZI-PrEP participants and other PrEP users. EZI-PrEP participants were all individuals enrolled in the EZI-PrEP study. The design and procedures of the EZI-PrEP study have been described previously^20^. In brief, participants were recruited by trained nurses during regular PrEP monitoring visits and from the waiting lists for enrolment in the NPP, at one of the four participating SHCs of Public Health Services in Amsterdam, Rotterdam, The Hague, and Nijmegen. The number of participants included by each SHC was roughly proportional to each centre’s maximum number, set by the NPP. Eligible individuals: (1) were 18 years or older; (2) lived in the catchment area of a participating sexual health centre; (3) were able to complete informed consent, medical history, a daily diary, and questionnaires in English or Dutch; (4) had an email address; (5) owned a smartphone capable of running the study application; (6) had daily access to an internet connection; and (7) being able to complete online bank transactions. Individuals were excluded from participation in the EZI-PrEP study in case of an HIV diagnosis or when, based on assessments by the study nurse or physician, online-mediated or 6-monthly monitoring would be inappropriate due to medical or psychosocial conditions^20^. After the PrEP consultation and explanation of the study, participants provided signed informed consent and were randomized to one of four study arms: (1) in-clinic monitoring every 3 months, (2) in-clinic monitoring every 6 months, (3) online monitoring every 3 months, or (4) online monitoring every 6 months. EZI-PrEP participants were requested to complete an online baseline questionnaire including questions about income level, choice of PrEP regimen, mental well-being through the Patient Health Questionnaire-9 (PHQ-9)^25^, sexual compulsivity scale (SCS)^26,27^, Alcohol Use Disorders Identification Test (AUDIT)^28–30^, and Drug Use Disorders Identification Test (DUDIT)^31,32^. For the comparison with other PrEP users, we used only the routinely collected data from the PrEP consultation as part of each participants’ inclusion visit.

Other PrEP users were individuals who were enrolled in the NPP in one of the four SHCs that participated in EZI-PrEP, did not opt-out of the use of their data, and did not participate in the EZI-PrEP study. Cisgender women and individuals <18 years old who were seeking PrEP care at one of the SHCs were excluded from analyses. We used the routinely collected data of the first PrEP consultation visit of each individual other PrEP user between 21 September 2021 and 9 August 2022 (i.e., the inclusion period of EZI-PrEP trial) (Textbox 1).

### 2.3 Variables

The analyses included variables on socio-demographic characteristics (e.g., age, gender, country of birth and highest completed or current educational level), sexual behaviour in the preceding 6 months (e.g., number of sexual partners, condom use, group sex, sex work, and use of alcohol and drugs during sex) and PrEP use in the preceding 3-6 months. Chemsex was defined as use of crystal methamphetamine, gamma-hydroxybutyrate (GHB)/gamma-butyrolactone (GBL), mephedrone or ketamine around the time of sex^33^. Chlamydia and gonorrhoeae infections were defined as a positive test result in at least one anatomical location; while infectious syphilis was defined as being diagnosed with syphilis I, syphilis II, or syphilis latens recens.

Only for EZI-PrEP participants, a PHQ-9 score ≥5 was used to indicate symptoms of depression^25^, and an SCS score ≥24 was used to indicate a greater impact of sexual thoughts on daily functioning and an inability to control sexual thoughts or behaviours^26,27^. Based on the AUDIT scores, we categorized alcohol use as follows: no or low-risk alcohol consumption (0-7), hazardous alcohol consumption (8-15), harmful alcohol consumption (16-19), and likely dependent on alcohol (≥20) ^28–30,34^. Based on the DUDIT scores we created three categories: no drug use related problems (≤5), harmful drug use (6-24), and likely dependent on drugs (≥25) ^31,32^.

### 2.4 Statistical analysis

Characteristics were compared (1) between SHCs overall (in only EZI-PrEP participants) and (2) between EZI-PrEP participants and other PrEP users using Wilcoxon rank-sum tests for continuous variables, and Pearson’s χ^2^ or Fisher’s exact for categorical variables. These analyses were also conducted per SHC.

All analyses were performed using STATA (v17.0, STATA Corporation, College Station, TX, USA).

### 2.5 Ethical considerations

The EZI-PrEP Trial was approved by the medical ethics committee of the Amsterdam University Medical Centre, file number 2020_154 - NL74494.018.20. The trial has been registered with ClinicalTrials.gov, trial number NCT05093036.

## 3. Results

### 3.1. Baseline characteristics of EZI-PrEP participants

Between 21 September 2021 and 9 August 2022, 469 EZI-PrEP participants were included: 464 cisgender males and five individuals identifying as transgender or gender diverse (Table 2).

Median age was 36 years [interquartile (IQR)=29-47], with 48 (10%) EZI-PrEP participants being <25 years old. Most participants (68%, n=319) were born in the Netherlands, 377 (82%) completed or pursued a university or college degree, and 408 (87%) had used PrEP in the 3 months preceding enrolment. Twelve participants (3%) had no formal education or only a primary education. The median number of sex partners in the 6 months before enrolment was 7 (IQR=4-15). During the same period, 447 (95%) EZI-PrEP participants reported condomless anal sex, 168 (38%) group sex, 123 (26%) chemsex, and 3 (1%) sex work. 84 (19%) participants were diagnosed with at least one bacterial STI at enrolment: *Neisseria gonorrhoea* infection, n=45 (10%); *Chlamydia trachomatis* infection, n=40 (9%); infectious syphilis, n=8 (2%).

Almost half (n=203, 47%) of EZI-PrEP participants, scored five or higher on PHQ-9, suggesting symptoms of depression. Few EZI-PrEP participants were likely dependent on alcohol (n=12, 3%) or drug use (n=7, 2%).

### 3.2. Comparing EZI-PrEP participants across sites

252 (54%) participants were included in Amsterdam, 112 (24%) in Rotterdam, 75 (16%) in The Hague, and 30 (6%) in Nijmegen (Table 2). Figure 1 visualizes the distribution of socio-demographic, sexual behaviour and mental health characteristics, as well as the prevalence of bacterial STIs, across these four public health services.

**Figure 1:**
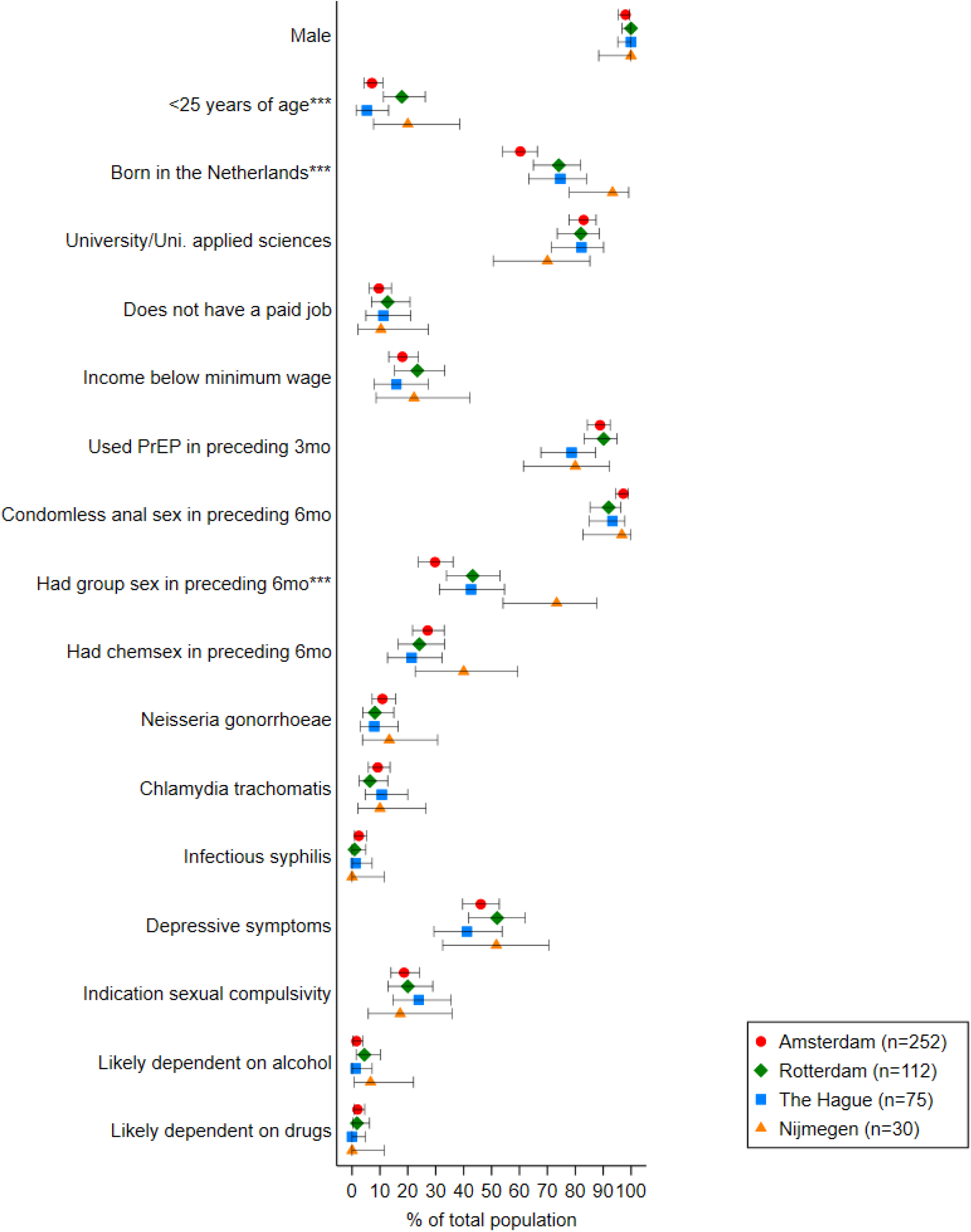
Baseline socio-demographic characteristics, sexual behaviour, STI prevalence and mental health of participants by Sexual Health Centre, EZI-PrEP study, the Netherlands, September 2021 - August 2022. Percentages are represented by markers, defined in the legend, and lines represent 95% Exact confidence intervals. For all comparisons p>0.05 except for *** p-value <0.001.Percentages are based on complete data; those with missing/unknown data were excluded from analysis. Number of missings can be found in Table 2. Abbreviations: EZI-PrEP: E-Health for Zero Infections - facilitating access to and use of Pre-Exposure Prophylaxis in the Netherlands; PrEP: pre-exposure prophylaxis; STI: sexually transmitted infection; mo: months.

The distribution of age differed between sites. Rotterdam (18%, 20/112) and Nijmegen (20%, 6/30) included a higher proportion of participants between 18 and 25 years of age than Amsterdam (7%, 18/252) and The Hague (5%, 6/75) (p=0.004). Also, the proportion born in the Netherlands was much lower in Amsterdam (60%, 152/252) and higher in Nijmegen (93%, 28/30) compared to Rotterdam (74%, 83/112) and the Hague (75%, 56/75) (p<0.001). Education level did not differ between the sites.

Regarding sexual behaviour in the 6 months before baseline measurement, we found no significant differences between the sexual health centres in the median number of sex partners, and the percentage of participants reporting insertive anal sex, condomless anal sex, receptive condomless anal sex, chemsex and injecting drugs around or during sex in the 6 months before baseline. However, the proportion of participants who reported receptive anal sex varied substantially between SHCs: 81% (205/252) in Amsterdam, 89% (100/112), in Rotterdam, 93% (70/75) in the Hague, and 83% (25/30) in Nijmegen (p=0.037). Moreover, 88% (223/252) of participants in Amsterdam reported condomless insertive anal sex in comparison to the other SHCs (respectively 88/112, 59/75, 24/30) (p=0.042). The percentage of participants who reported group sex was 43% in Rotterdam and the Hague, while higher in Nijmegen (73%, 22/30) and lower in Amsterdam (30%, 66/222), p<0.001, Figure 1).

### 3.3. Comparisons of EZI-PrEP participants with other PrEP users

We included 469 EZI-PrEP participants (464 MSM and 5 transgender en gender diverse persons) and 5,161 (4954 MSM and 207 transgender en gender diverse persons) other PrEP users of the NPP (Table 1). Figure 2 visualizes the distribution of socio-demographic and sexual behaviour characteristics and STI prevalence of EZI-PrEP and other PrEP users.

**Figure 2:**
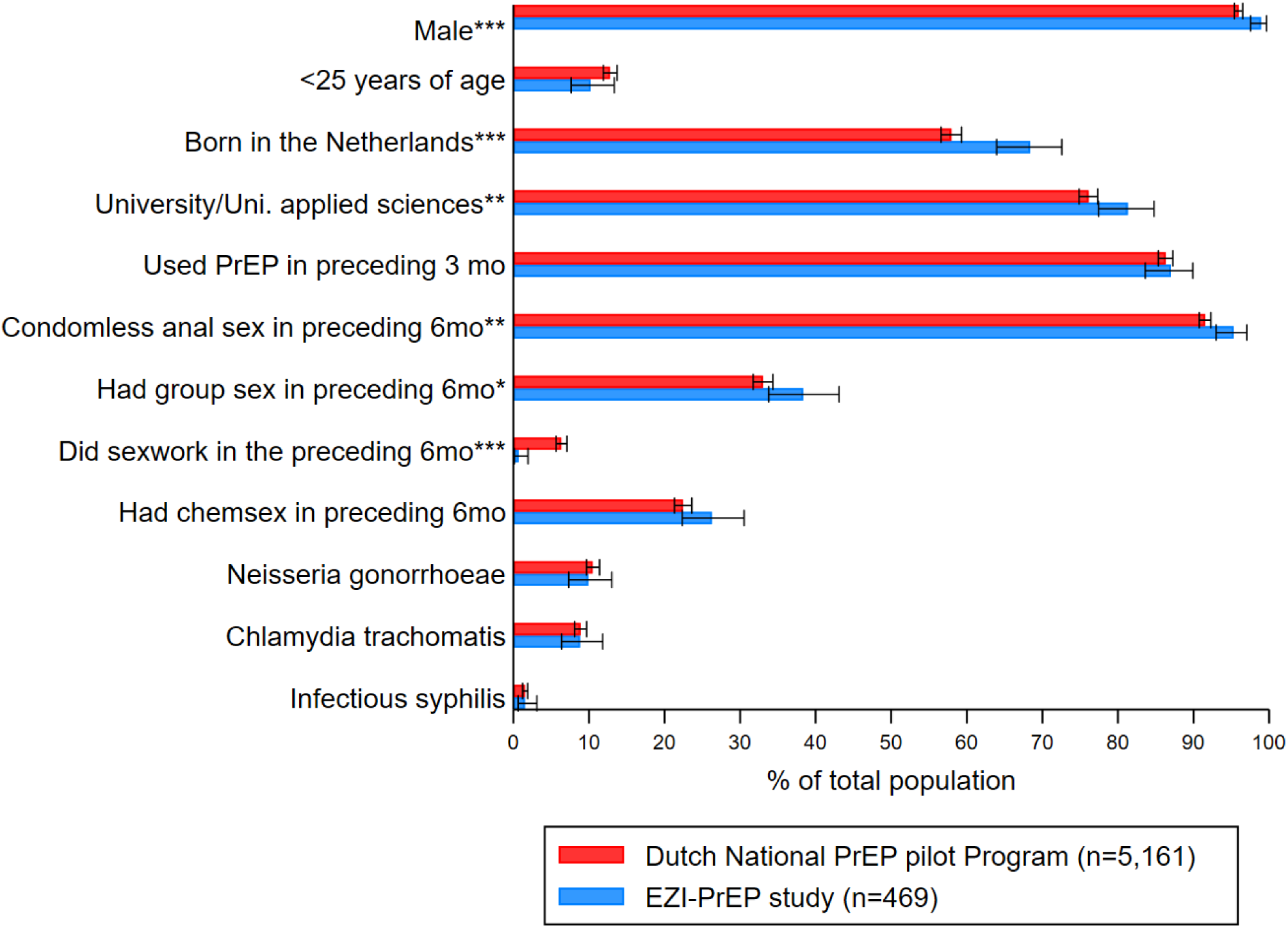
Baseline socio-demographic characteristics and sexual behaviour of EZI-PrEP participants and other PrEP users of the Dutch National PrEP pilot Programme in Sexual Health Centres in Amsterdam, Rotterdam, the Hague and Nijmegen, September 2021 - August 2022. Percentages are represented by markers, defined in the legend, and lines represent 95% Exact confidence intervals. * p-value < 0.05, ** p-value < 0.01, *** p-value < 0.001. Notes: Percentages are based on complete data; missing/unknown data excluded from calculations. Number of missings can be found in Supplementary Table 1. Abbreviations: EZI-PrEP: E-Health for Zero Infections -facilitating access to and use of Pre-Exposure Prophylaxis in the Netherlands; PrEP: pre-exposure prophylaxis.

**Table 1.**
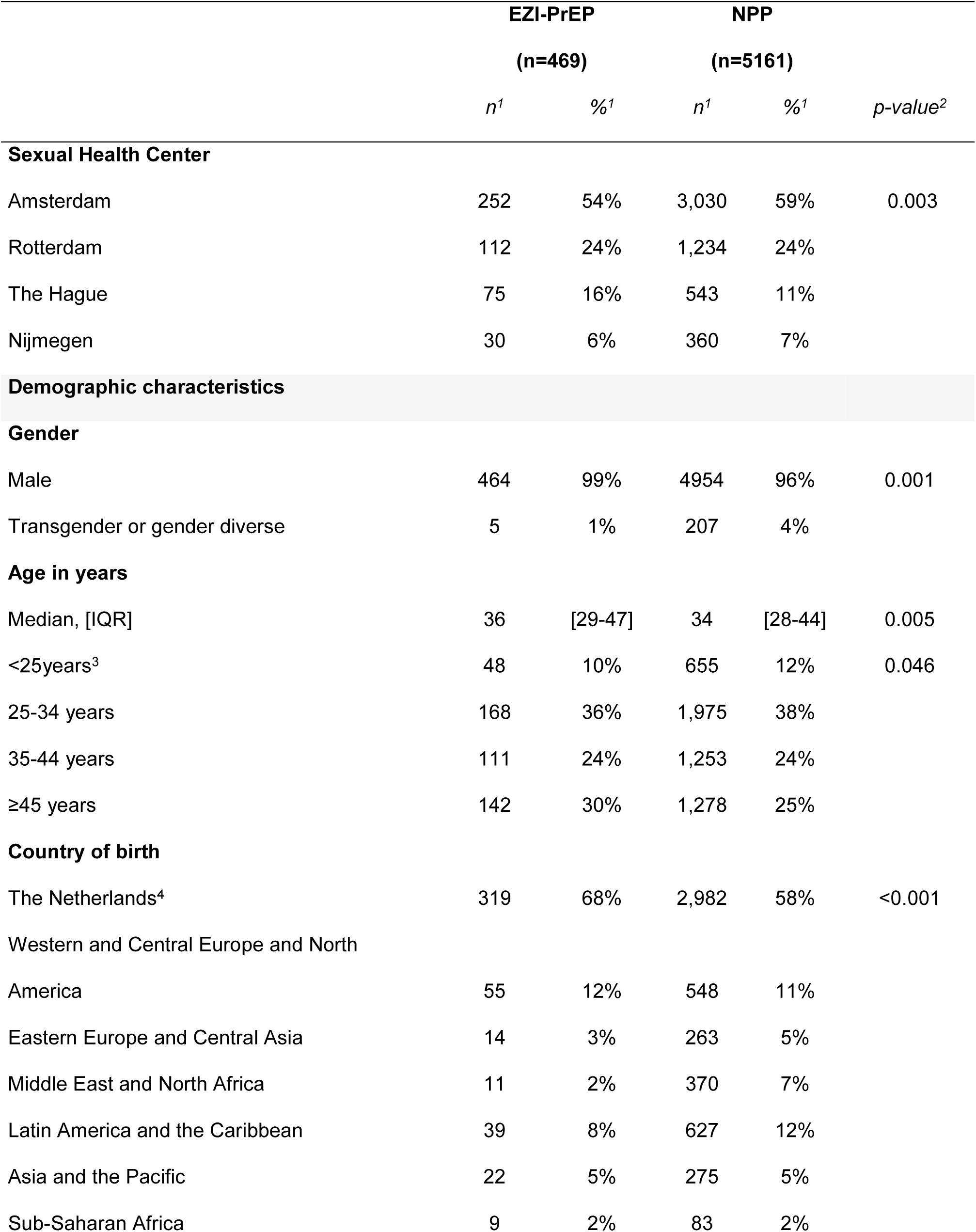
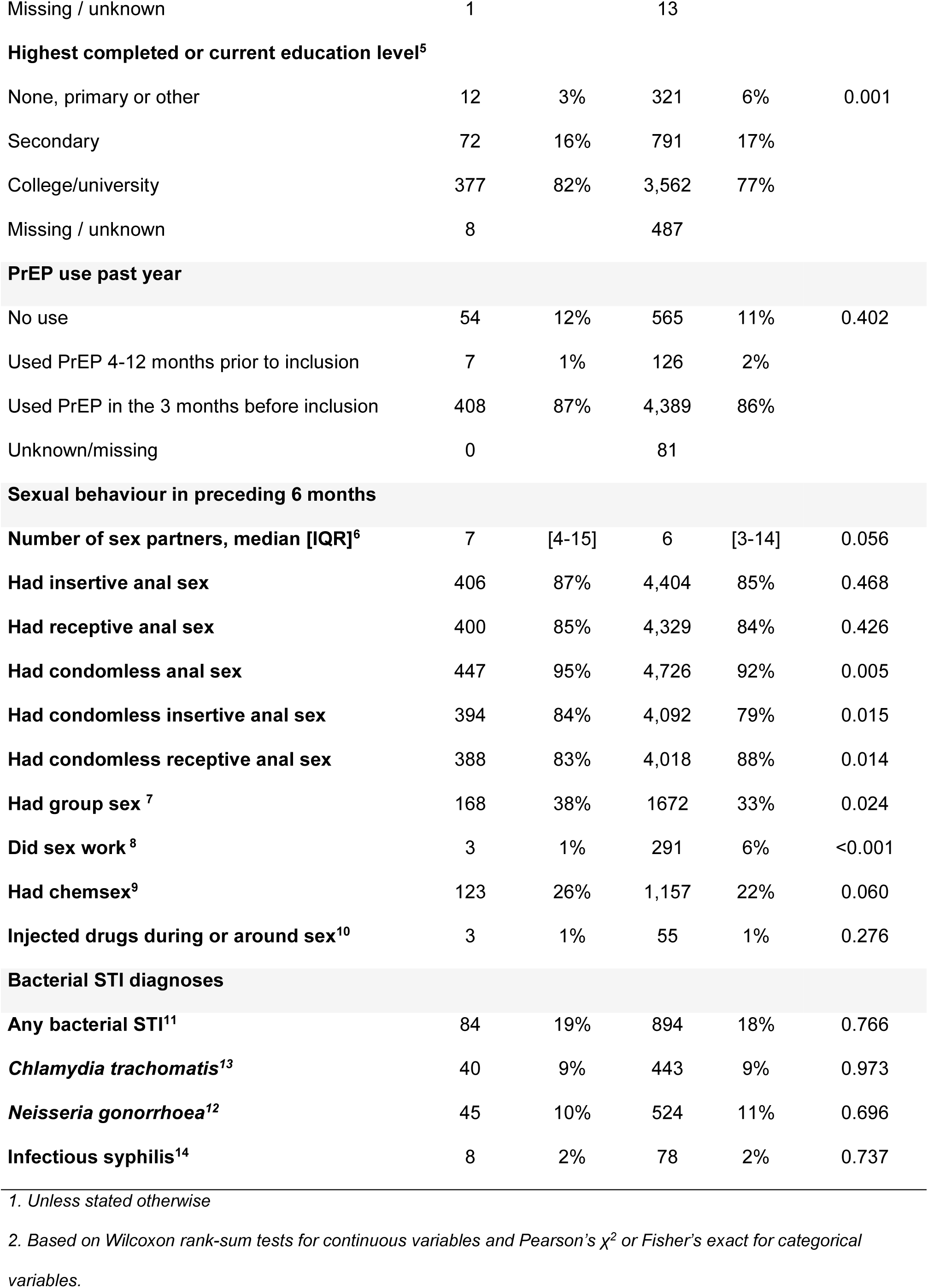
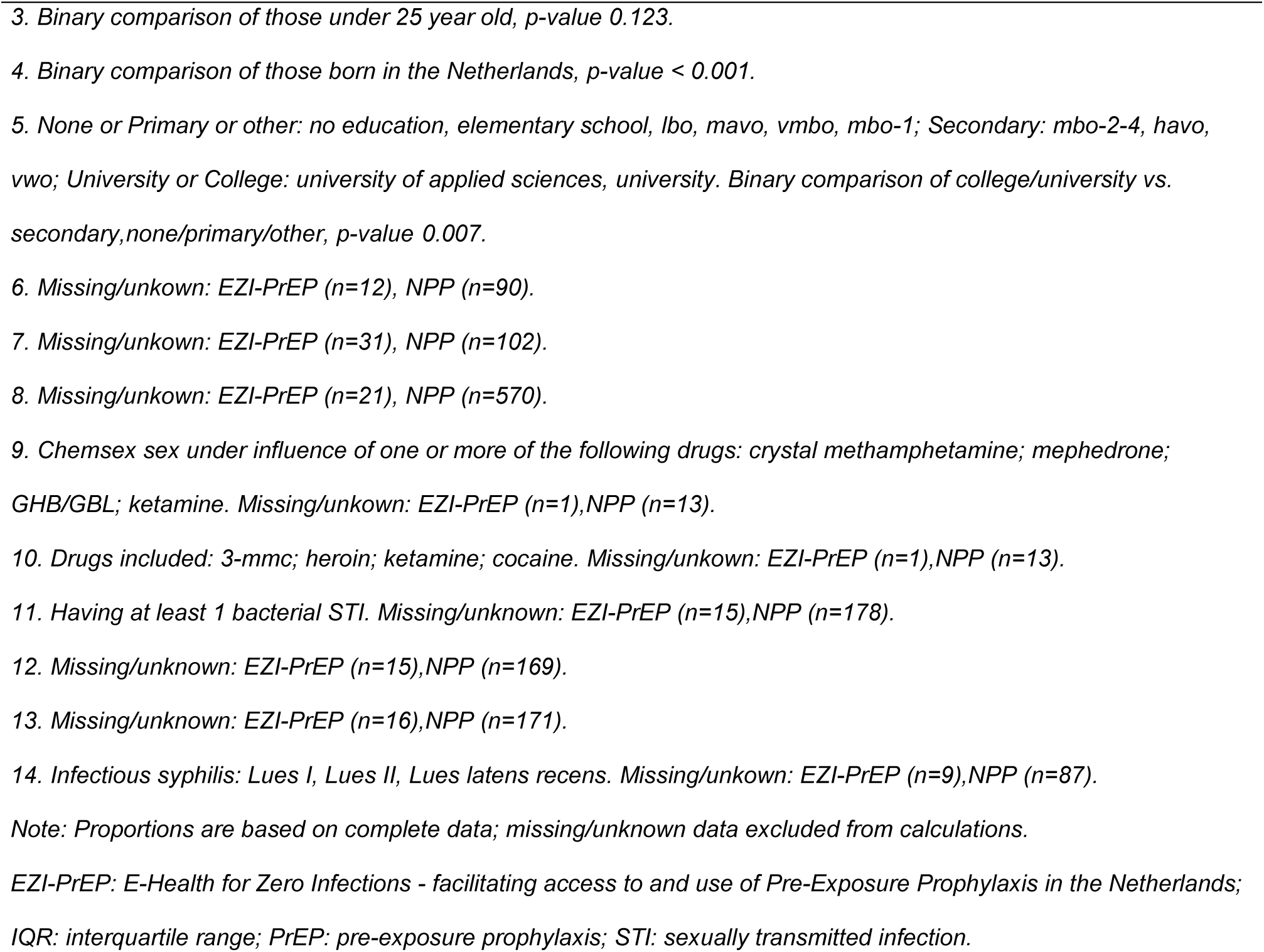
Baseline socio-demographic characteristics and sexual behaviour of EZI-PrEP participants and other PrEP users of the Dutch National PrEP pilot Programme at the Sexual Health Centres in Amsterdam, Rotterdam, The Hague and Nijmegen, September 2021 to August 2022

**Table 2.**
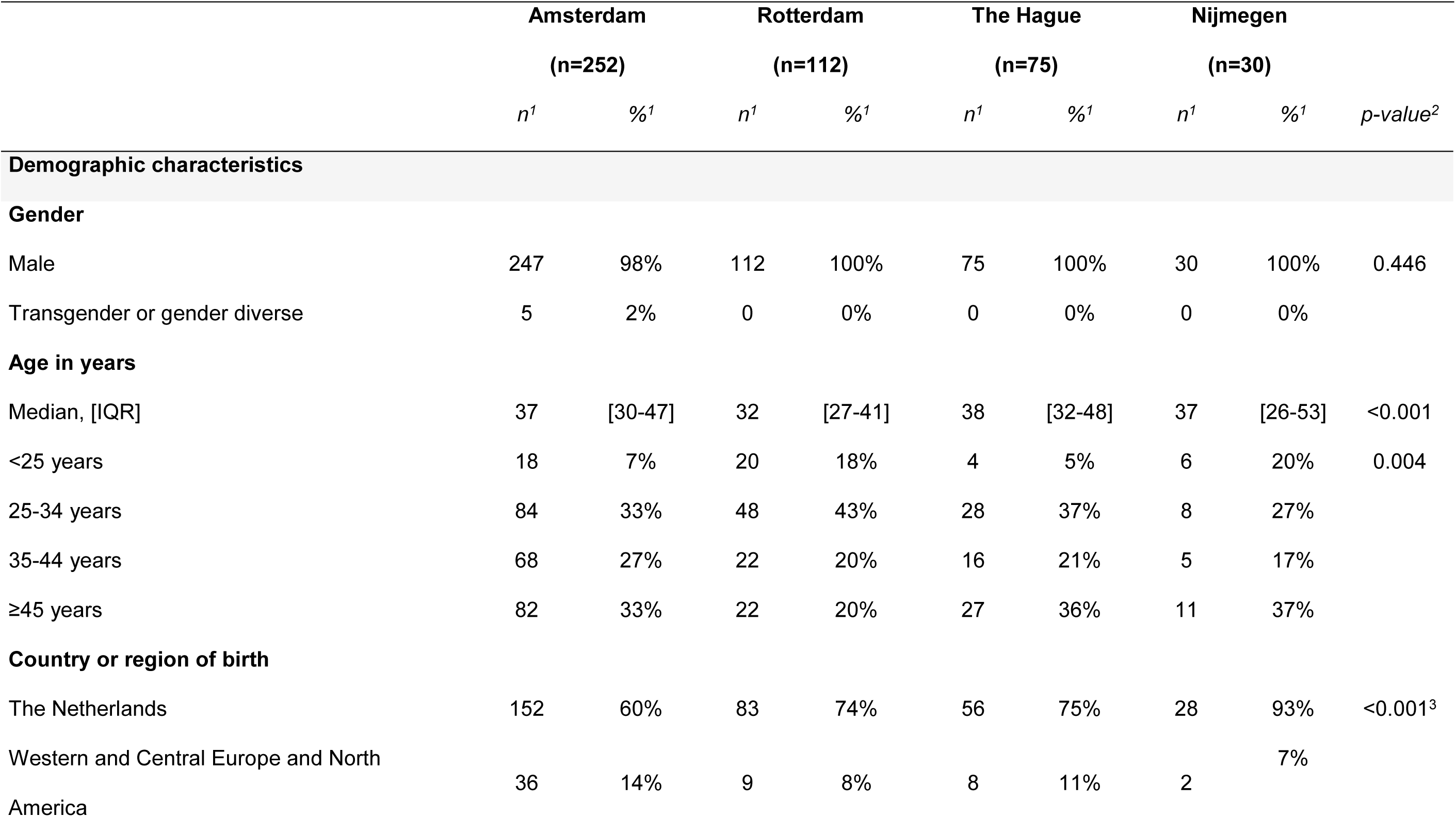
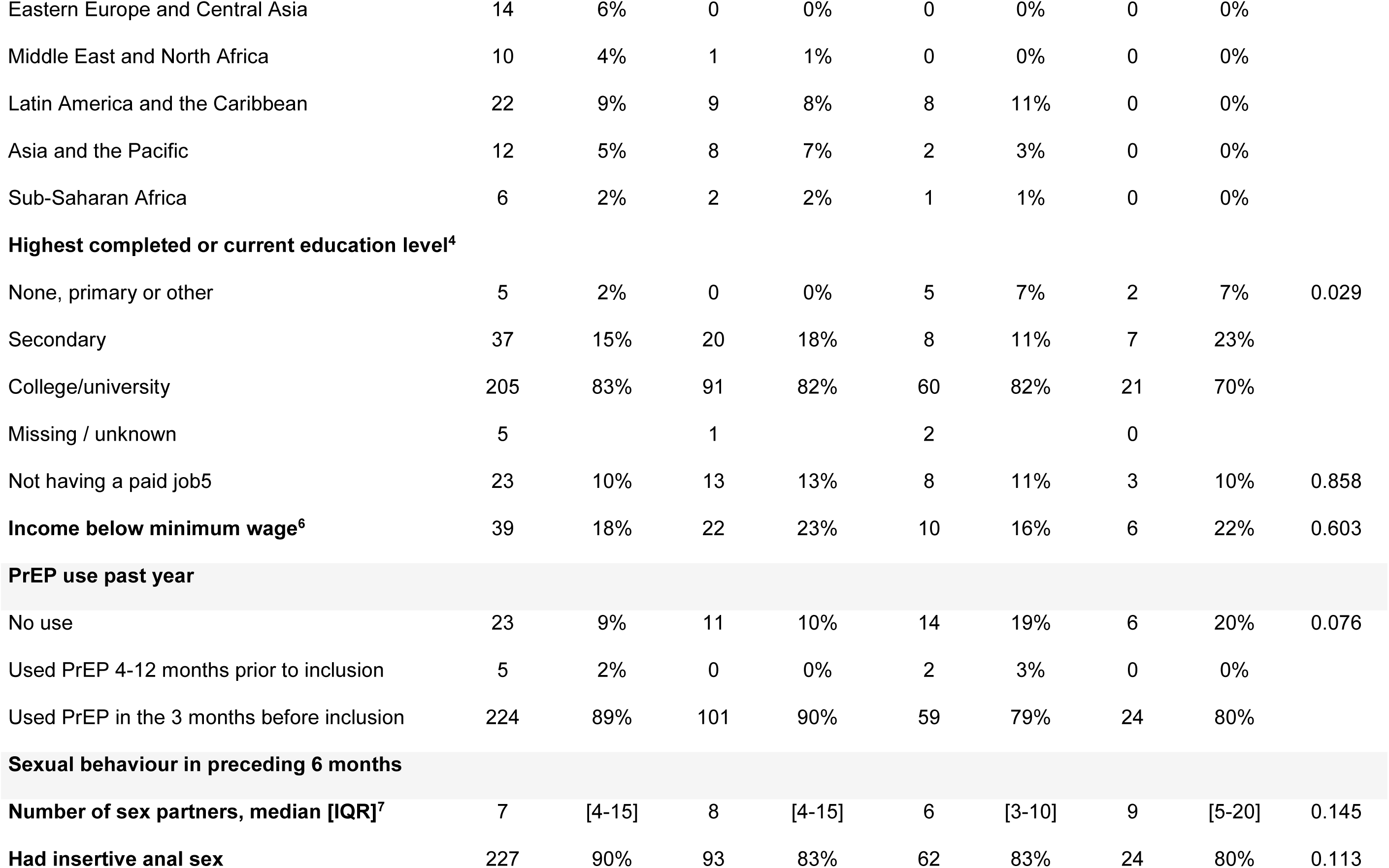
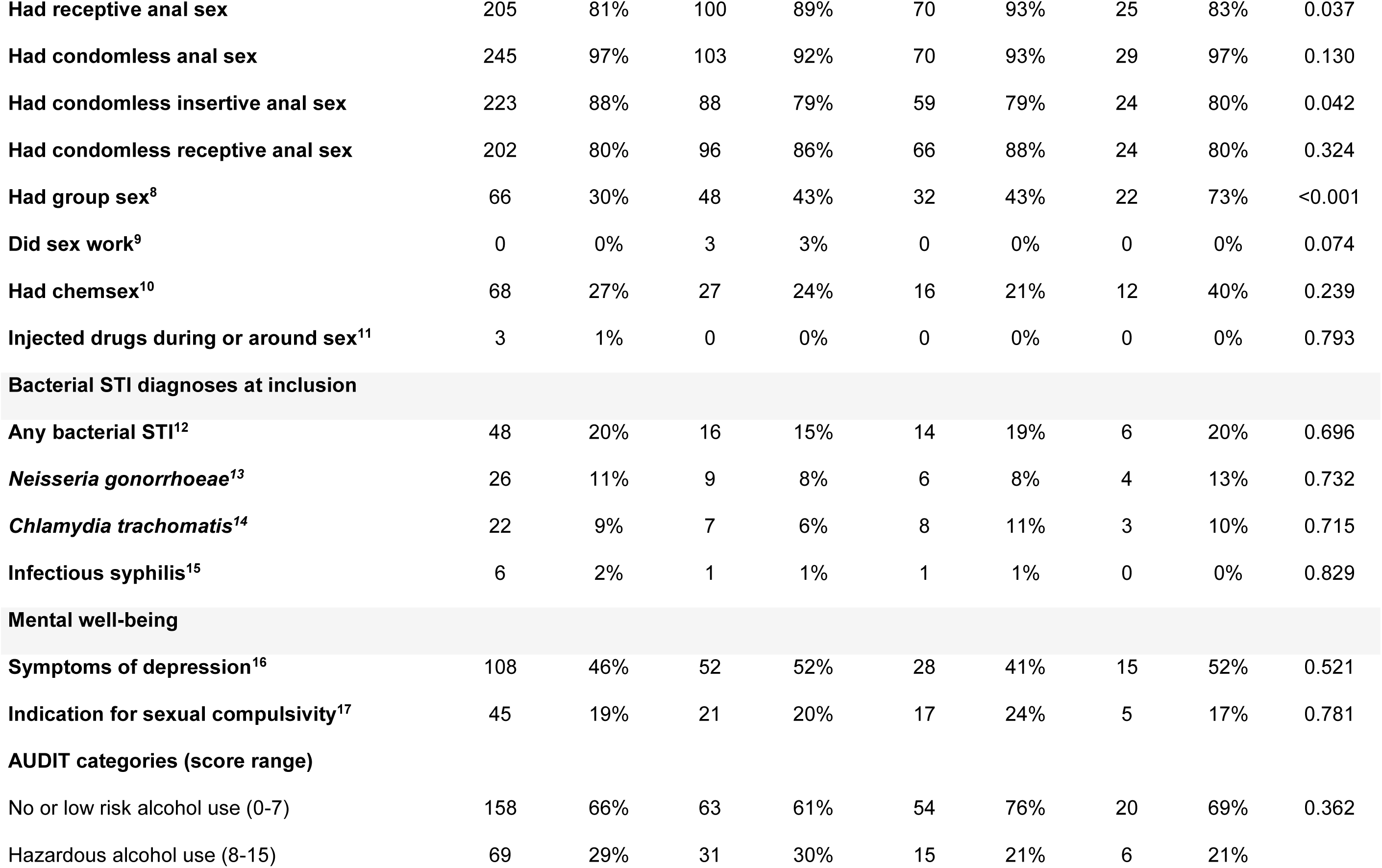
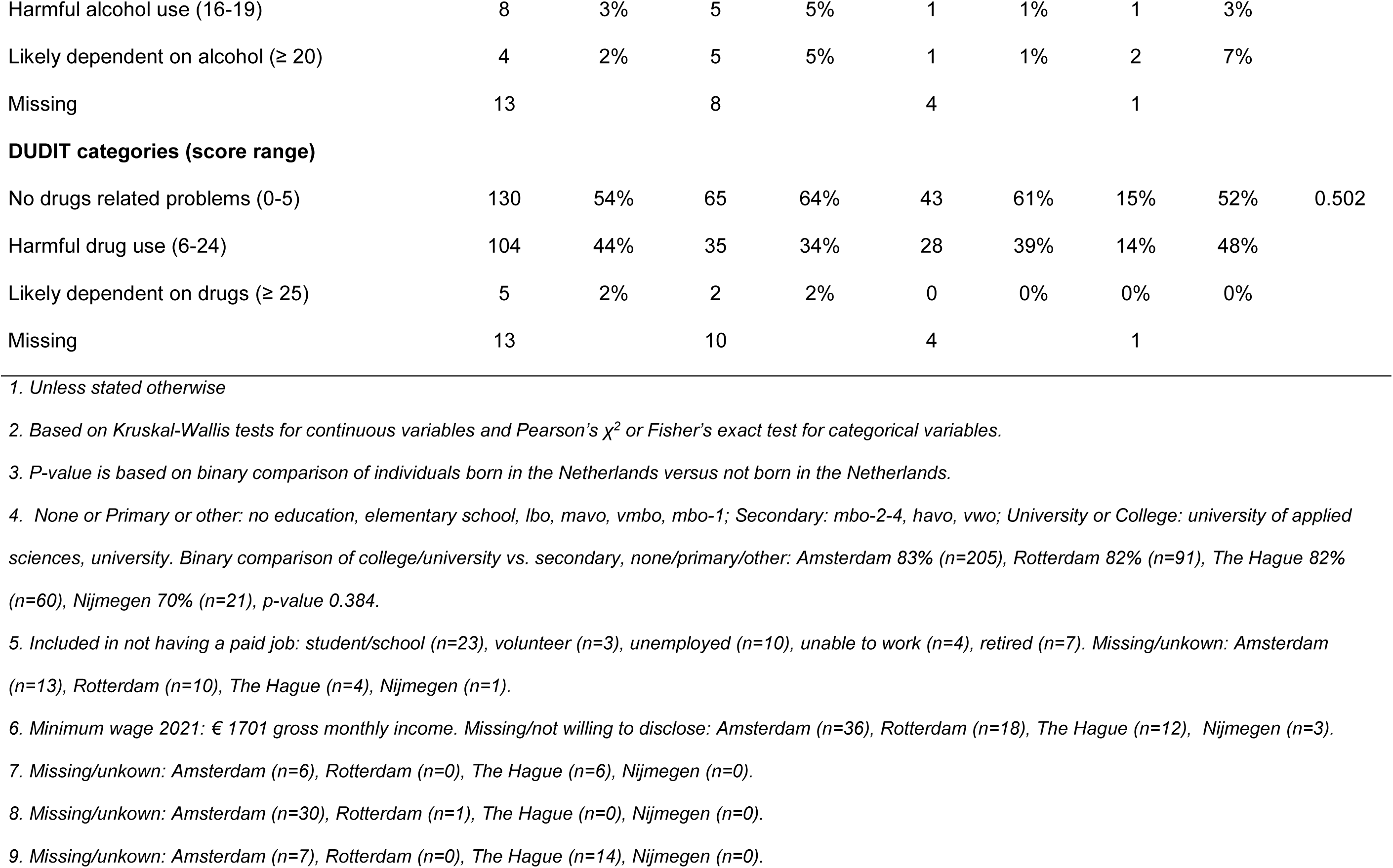
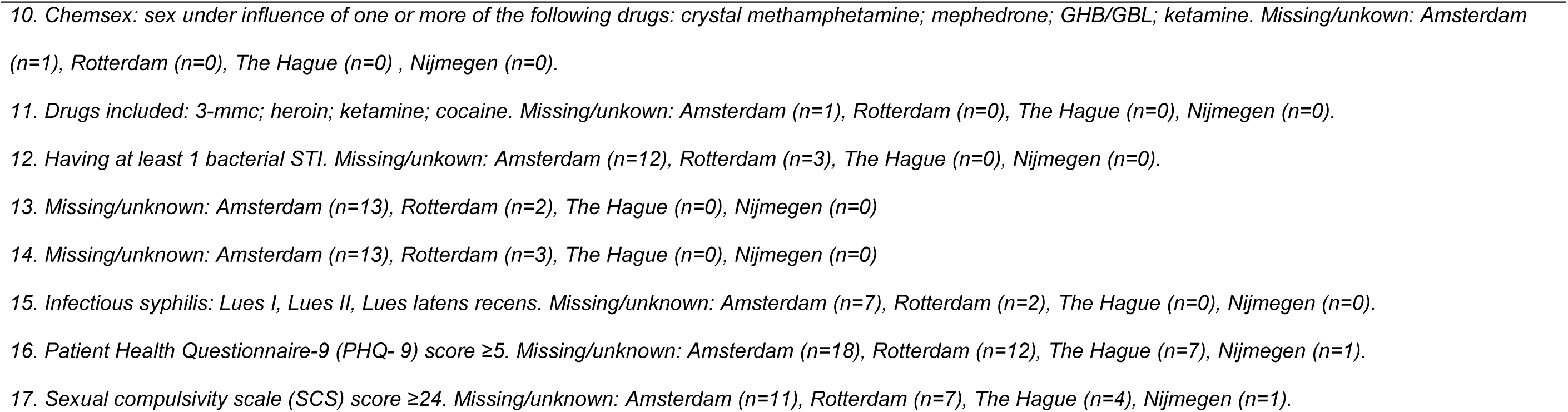
Baseline socio-demographic characteristics, sexual behaviour, STI prevalence and mental health of participants by Sexual Health Centre, EZI-PrEP study, the Netherlands, September 2021 - August 2022

EZI-PrEP participants were more often male than the other PrEP users (99% vs. 96% p<0.001), more often born in the Netherlands (68% vs. 58%, p<0.001), and more often completed or were pursuing a college or university degree (82% vs. 77%, p=0.007). EZI-PrEP participants were slightly older (median=36 years, IQR=29-47) than other PrEP users (34 years, IQR=28-44) (p=0.005), but the proportion of individuals below 25 years old was not significantly different (10% and 12% respectively, p=0.12).

With respect to sexual behaviour in the preceding 6 months, we found no significant differences between the two groups in the distribution of the number of sex partners, the percentage participants reporting insertive or receptive anal sex, chemsex and injecting drug use during or around sex in the previous 6 months. However, EZI-PrEP participants more often reported condomless anal sex (95% vs. 92%, p=0.005) and group sex (38% vs. 33%, p=0.024), but less often reported condomless receptive anal sex (83% vs. 88%, p=0.014) and sex work (1% vs. 6%, p<0.001). The overall prevalence of any bacterial STIs was similar in both groups (19% vs. 18%, p=0.799). Also the prevalence of each individual STI was similar in both groups, i.e., *Chlamydia trachomatis* (9% vs. 9%, p=0.973), *Neisseria gonorrhoea* (10% vs. 11%, p=0.696), infectious syphilis (2% vs. 2%, p=0.737)

### 3.4. Comparing EZI-PrEP participants and other PrEP users per sexual health centre

With respect to socio-demographic variables, we found that EZI-PrEP participants in The Hague and Nijmegen were largely similar to the other PrEP users in the respective sexual health centres, although the median age of EZI-PrEP participants in The Hague was higher (38 years [IQR 32-48]) than the other PrEP users in The Hague (34 years [IQR 27-46], p=0.012) (Supplementary Tables 1c-d). EZI-PrEP participants in Rotterdam had a similar age to the other PrEP users in Rotterdam and more often completed or were pursuing a college or university degree (82% vs. 72%, p=0.023) (Supplementary Table 1b). EZI-PrEP participants in Amsterdam were older (median=37 years [IQR 30-47]) than the other PrEP users in Amsterdam (33 years [IQR 28-43], p<0.001) and were more often born in the Netherlands (60% vs. 51%, p=0.013). However, a similar proportion of EZI-PrEP participants and other PrEP users in Amsterdam completed or were pursuing a college or university degree (83% vs. 81%, p=0.354) (Supplementary Table 1a).

With respect to sexual behaviour in the preceding 6 months, we found no significant differences between EZI-PrEP participants and other PrEP users in Rotterdam and The Hague. Overall, EZI-PrEP participants in Nijmegen reported similar sexual behaviour as the other PrEP users in Nijmegen. However, EZI-PrEP participants in Nijmegen reported a higher median number of sex partners (median=9 [IQR 5-20]) than other PrEP users in Nijmegen (median=5 [IQR 3-10], p=0.013) and more often reported group sex (73% vs. 50%, p=0.013). For Amsterdam, we found no significant differences between the two populations in the distribution of the number of sex partners, group sex, and the percentage of participants reporting receptive anal sex, condomless receptive anal sex, chemsex and injecting drug use during or around sex, in the past 6 months. However, EZI-PrEP participants more often reported insertive anal sex (90% vs. 85%, p=0.038), condomless anal sex (97% vs. 91%, p<0.001) and condomless insertive anal sex (88% vs. 79%, p<0.001), but less often sex work (0% vs. 10%, p<0.001) than the other PrEP users in Amsterdam. The prevalence of bacterial STIs was similar in both populations across all four sexual health centres.

## 4. Discussion

In this study, we compared EZI-PrEP participants with other PrEP users in the NPP to assess representativeness of the EZI-PrEP study participants. We did observe some notable differences in the characteristics between the two populations, of which the most important is that EZI-PrEP participants were less often transgender or gender-diverse, slightly older, more often born in the Netherlands, and more often completed or were pursuing a university or college degree. Furthermore, we found that EZI-PrEP participants reported sexual behaviours associated with STI acquisition more often, but reported sex work less often. Notwithstanding these differences, we observed importantly that the STI prevalence rates were similar in the two populations.

There were noteworthy differences in sexual behaviours between EZI-PrEP participants and other PrEP users (i.e., more group sex and condomless anal sex). The numerical differences were nonetheless small and were unlikely to be of much clinical relevance. Indeed, there was no difference in STI prevalence between groups. When comparing the STI prevalence in the EZI-PrEP and NPP populations in our study to national STI-positivity rates among MSM PrEP users in 2022, we found similar rates (i.e., gonorrhoea 9.2%, chlamydia 9.4%, infectious syphilis 1.7%)^35^. This concordance with national data further supports our observation that STI rates among EZI-PrEP participants closely reflect those observed in the broader MSM population using PrEP. In studies of key populations where PrEP is not being used, the presence of an STI is highly associated with HIV and hence is considered to be an adequate proxy for the risk of HIV acquisition^36,37^. Given the similar prevalence of STI between populations in our analyses, as well as the similarity with national rates, the risk of acquiring HIV among EZI-PrEP participants could be considered generalizable to the broader population of PrEP users in the Netherlands.

Certain groups were clearly underrepresented in the EZI-PrEP study. Transgender and gender-diverse individuals comprised only 1% of EZI-PrEP participants compared to 4% of the other PrEP users in the NPP. Furthermore, PrEP users who were not born in the Netherlands were less commonly enrolled in the EZI-PrEP trial. Prior research has suggested that transgender and gender-diverse persons, as well as recently migrated MSM, face specific barriers to HIV and PrEP care and a higher risk of HIV acquisition^38–42^. Individuals who completed no or primary schooling were less often enrolled in the EZI-PrEP trial compared to other PrEP users. While the level of education may not necessarily directly influence the risk of HIV acquisition, it can affect the effective use of telehealth services, potentially increasing HIV acquisition risk due to lower technological or medical literacy ^43–46^. For this group, in-clinic PrEP care may be more suitable. Finally, sex work was less commonly reported among EZI-PrEP participants, while sex workers are known to belong to a key population at a higher risk of HIV acquisition ^47–50^. Taken together, some caution is warranted when extrapolating the findings from the EZI-PrEP trial to these underrepresented groups. Tailored PrEP care would be essential to meet the specific needs of these populations.

Within the EZI-PrEP population, comparisons across sites revealed that participants at the four study sites were for the most part comparable. Notable differences included the younger age of participants in Rotterdam and a higher percentage of participants who reported group sex in Nijmegen. In parallel, when comparing the EZI-PrEP participants in Rotterdam and Nijmegen to their respective populations of other PrEP users in the NPP, we observed the same differences in socio-demographics and sexual behaviour. These differences may suggest a selection effect, with certain groups being over- or underrepresented due to site-specific factors. Given the small sample size in Nijmegen (n=30), such selection effects could be more pronounced, potentially limiting the generalizability of the findings from this site. However, considering that the STI prevalence was similar between EZI-PrEP participants and other PrEP users within centres, none of these differences in socio-demographics or sexual behaviours appeared to have had any clinically meaningful effect.

There are certain limitations worth mentioning. The EZI-PrEP study did not include individuals who may be economically or socially vulnerable, such as those without a bank account, postal address, or the ability to complete online transactions. This limitation was unavoidable due to the online-mediated PrEP monitoring being examined. Consequently, homeless and undocumented individuals, as well as those with limited digital skills, were not included in the study. Additionally, we did not compare EZI-PrEP participants with individuals receiving PrEP through their general practitioner, which may be a distinct group of PrEP users. Furthermore, considering the large number of individuals in the NPP, many of the comparisons between EZI-PrEP participants and other PrEP users were likely to have been overpowered and some of the significant findings are unlikely to bear any clinically meaningful difference.

In conclusion, the characteristics of EZI-PrEP participants are largely congruent to those of other PrEP users in the NPP, and hence the results of the EZI-PrEP trial could be considered broadly generalizable to the wider population of PrEP users. Nevertheless, the underrepresentation of transgender and gender-diverse persons, individuals born outside the Netherlands, sex workers and people with no or primary education do pose an issue with generalizability. Research beyond EZI-PrEP may need to focus on delivering other PrEP modalities for these underrepresented populations.

## Authors contributions

UD, MFSvdL, AB, VJ, EH, MP, HMG, FvH, and MLGB contributed to study conceptualisation and design. MLGB, UD, MFSvdL, LB, CvB, JW, SB, MS, HMG, LW and MSA contributed to data acquisition and data management. MLGB, FD, KvdV, MFSvdL, UD, and VJ contributed to analysis and interpretation of the data. MLGB drafted the manuscript. All authors contributed to critical revision and approved the final manuscript.

## Acknowledgements

We would like to thank our study participants for their time and input. We would also like to thank the nurses and physicians involved in the EZI-PrEP trial: Kenneth Yap, Dennis Heineman, Francine van den Heuvel, Ilya Peters, Vhi Nhi Nguyen, Abdullah Aksoy, Maudy Sluimer, Marijn Stip, Charelle Zwiggelaar, Jean-Marie Brand, Emma van de Ven, Alexander Sangers, and Daphne Sassen. Additionally we thank others who have contributed to design, data collection, data management or logistics of the EZI-PrEP study: Janneke Bil, Eline Wijstma, Mark van den Elshout, Liza Coyer, Amy Matser, Liesbeth Flobbe, Frank Mouthaan, Ertan Ersan, Dominique Loomans, Maarten van Haagen, Karin Tang, Marjolein Booij, Bart-Jan Mulder, Joël Illidge, Roelien Prins, Arjan van Beijnen, Wim Zuilhof, Noëmie Nijsten, Ilya Peters, Sarah Brevet, Jeannine Hautvast, Amber Hof, Remy Welleman, Sophie Kuizenga Wessel, Fokke Postma and Fetzen de Groot. Furthermore we would like to thank colleagues at the laboratories: Margreet Visser, René Vork, Wilma Vermeulen-Oost, Sylvia Bruisten at the Amsterdam regional microbiology laboratory; Mireille van Westreenen and colleagues at the department of Medical Microbiology at Erasmus MC laboratory and Annemiek Baltissen-van der Eijk and colleagues of the virology department; Suzanne van Veen and colleagues at Haaglanden Medical Centre; Matthew McCall and colleagues of the department of Medical Microbiology at Radboud UMC. Also, many thanks to Rosalie Zieck and Laurens Maussen from DC Pharmacy. Finally, we would like to thank the members of the EZI-PrEP advisory group for their feedback on design, data-collection and analyses of the EZI-PrEP study: Bea Vuylsteke, Marc van der Valk, Chris Tearno, David Burger, Hanna Bos, Jan den Hollander, John de Wit, Janny Dekker, Kai Jonas, Silke David, Sebastiaan Verboeket, Janneke van de Wijgert, and Paul Zantkuijl.

## Data Availability Statement

The EZI-PrEP data are owned by the EZI-PrEP consortium. EZI-PrEP data can be requested by submitting a study proposal to the EZI-PrEP steering committee. It will be possible to obtain a proposal format from the Public Health Services of Amsterdam. Requests for further information can also be submitted through the same email addresses. The EZI-PrEP steering committee will verify each proposal for compatibility with general objectives, ethical approval, and informed consent forms of the EZI-PrEP trial and potential overlap with ongoing studies. There will be no restrictions to obtaining the data and all data requests will be processed in a similar way.

---

**Textbox 1.**
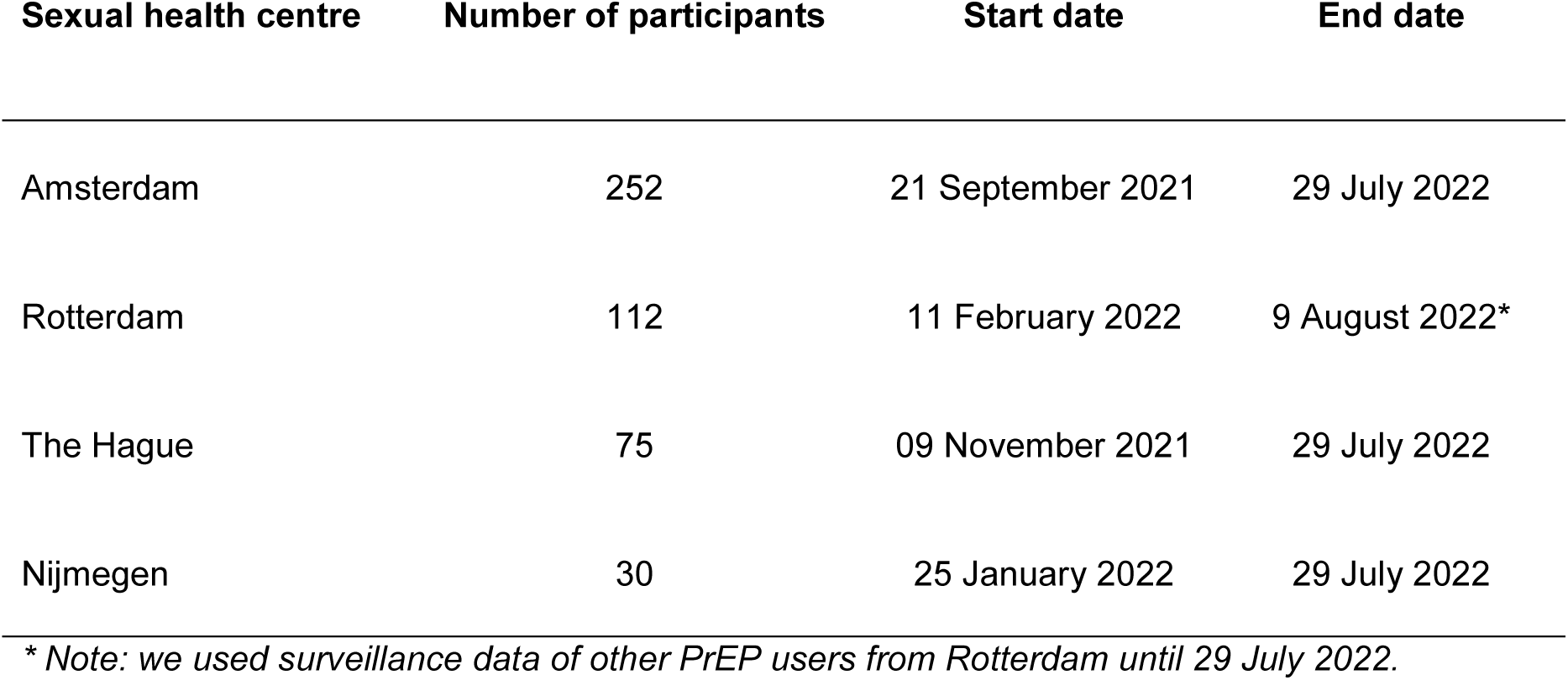
Overview of the number of participants, start and end date of the inclusion period, per sexual health centres.

| **Sexual health centre** | **Number of participants** | **Start date** | **End date** |
| --- | --- | --- | --- |
| Amsterdam | 252 | 21 September 2021 | 29 July 2022 |
| Rotterdam | 112 | 11 February 2022 | 9 August 2022* |
| The Hague | 75 | 09 November 2021 | 29 July 2022 |
| Nijmegen | 30 | 25 January 2022 | 29 July 2022 |

*\* Note: we used surveillance data of other PrEP users from Rotterdam until 29 July 2022.*

## References

1. Fonner VA, Dalglish SL, Kennedy CE, et al. Effectiveness and safety of oral HIV preexposure prophylaxis for all populations. AIDS 2016;30(12):1973–83.

2. Molina JM, Capitant C, Spire B, et al. On-Demand Preexposure Prophylaxis in Men at High Risk for HIV-1 Infection. N Engl J Med 2015;373(23):2237–46.

3. Molina JM, Charreau I, Spire B, et al. Efficacy, safety, and effect on sexual behaviour of on-demand pre-exposure prophylaxis for HIV in men who have sex with men: an observational cohort study. Lancet HIV 2017;4(9):e402–e410.

4. Grant RM, Lama JR, Anderson PL, et al. Preexposure chemoprophylaxis for HIV prevention in men who have sex with men. N Engl J Med 2010;363(27):2587–99.

5. McCormack S, Dunn DT, Desai M, et al. Pre-exposure prophylaxis to prevent the acquisition of HIV-1 infection (PROUD): effectiveness results from the pilot phase of a pragmatic open-label randomised trial. Lancet 2016;387(10013):53–60.

6. Zhang J, Li C, Xu J, et al. Discontinuation, suboptimal adherence, and reinitiation of oral HIV pre-exposure prophylaxis: a global systematic review and meta-analysis. Lancet HIV 2022;9(4):e254–e268.

7. Chou R, Evans C, Hoverman A, et al. Preexposure Prophylaxis for the Prevention of HIV Infection: Evidence Report and Systematic Review for the US Preventive Services Task Force. JAMA 2019;321(22):2214–2230.

8. Rotsaert A, Vanhamel J, Vanbaelen T, et al. HIV Pre-exposure Prophylaxis (PrEP) Care in Belgium: A mixed-methods Study on PrEP Users’ Experiences and Service Delivery Preferences. AIDS Behav 2023.

9. Bavinton BR, Grulich AE. HIV pre-exposure prophylaxis: scaling up for impact now and in the future. Lancet Public Health 2021;6(7):e528–e533.

10. Twisk DE, Meima B, Nieboer D, Richardus JH, Gotz HM. Distance as explanatory factor for sexual health centre utilization: an urban population-based study in the Netherlands. Eur J Public Health 2021;31(6):1241–1248.

11. Owens C, Moran K, Mongrella M, et al. “It’s Very Inconvenient for Me”: A Mixed-Method Study Assessing Barriers and Facilitators of Adolescent Sexual Minority Males Attending PrEP Follow-Up Appointments. AIDS Behav 2022;26(1):21–34.

12. Zimbile F, David S, Daemen M, et al. Introducing video consultations at public sexual health clinics in the Netherlands: a mixed-methods study. Health Promot Int 2022;37(5).

13. Edwards J. Could the use of digital services improve the provision of HIV PrEP in the UK? BMJ 2022;377:o1562.

14. Kohler S, Dalal S, Hettema A, et al. Out-of-pocket Expenses and Time Spent on Clinic Visits Among HIV Pre-exposure Prophylaxis Users and Other Clinic Attendees in Eswatini. AIDS Behav 2023;27(4):1222–1233.

15. Kularadhan V, Gan J, Chow EPF, Fairley CK, Ong JJ. HIV and STI Testing Preferences for Men Who Have Sex with Men in High-Income Countries: A Scoping Review. Int J Environ Res Public Health 2022;19(5).

16. Mayer KH, Agwu A, Malebranche D. Barriers to the Wider Use of Pre-exposure Prophylaxis in the United States: A Narrative Review. Adv Ther 2020;37(5):1778–1811.

17. Hillis A, Germain J, Hope V, McVeigh J, Van Hout MC. Pre-exposure Prophylaxis (PrEP) for HIV Prevention Among Men Who Have Sex with Men (MSM): A Scoping Review on PrEP Service Delivery and Programming. AIDS Behav 2020;24(11):3056–3070.

18. Ong JJ, Fu H, Baggaley RC, et al. Missed opportunities for sexually transmitted infections testing for HIV pre-exposure prophylaxis users: a systematic review. J Int AIDS Soc 2021;24(2):e25673.

19. Not PrEPared: Barriers to accessing HIV prevention drugs in England. November 2022.Link to report: file:///W:/Downloads/Barriers-to-PrEP-in-England.pdf Accessed Last accessed: 10 February 2023.

20. Groot Bruinderink ML, Boyd A, Coyer L, et al. Online-Mediated HIV Pre-exposure Prophylaxis Care and Reduced Monitoring Frequency for Men Who Have Sex With Men: Protocol for a Randomized Controlled Noninferiority Trial (EZI-PrEP Study). JMIR Res Protoc 2023;12:e51023.

21. Mindell J, Aresu M, Bécares L, Tolonen H. Representativeness of participants in a cross-sectional health survey by time of day and day of week of data collection. European Journal of Public Health 2011;22(3):364–369.

22. Stuart EA, Bradshaw CP, Leaf PJ. Assessing the generalizability of randomized trial results to target populations. Prev Sci 2015;16(3):475–85.

23. Rothwell PM. External validity of randomised controlled trials:“to whom do the results of this trial apply?”. The Lancet 2005;365(9453):82–93.

24. A.T. Urbanus, C. Blom, David S. Implementatie PrEP-verstrekking en medische begeleiding in Nederland. Rijksinstituut voor Volksgezondheid en Milieu. Available from: https://zoek.officielebekendmakingen.nl/blg-872235.pdf., 2018.

25. Kroenke K, Spitzer RL, Williams JB. The PHQ-9: validity of a brief depression severity measure. J Gen Intern Med 2001;16(9):606–13.

26. Kalichman SC, Johnson JR, Adair V, et al. Sexual sensation seeking: scale development and predicting AIDS-risk behavior among homosexually active men. J Pers Assess 1994;62(3):385–97.

27. Kalichman SC, Rompa D. The Sexual Compulsivity Scale: further development and use with HIV-positive persons. J Pers Assess 2001;76(3):379–95.

28. Reinert DF, Allen JP. The Alcohol Use Disorders Identification Test (AUDIT): a review of recent research. Alcohol Clin Exp Res 2002;26(2):272–9.

29. Reinert DF, Allen JP. The Alcohol Use Disorders Identification Test (AUDIT): A Review of Recent Research. Alcoholism: Clinical and Experimental Research 2006;26(2):272–279.

30. Babor TF H-BJ, Saunders JB, Monteiro, M.G. The alcohol use disorders identification test: guideline for use in primary care. Geneva: World Health Organization, 2001.

31. Berman AH, Bergman H, Palmstierna T, Schlyter F. Evaluation of the Drug Use Disorders Identification Test (DUDIT) in criminal justice and detoxification settings and in a Swedish population sample. European addiction research 2005;11(1):22–31.

32. Hildebrand M. The psychometric properties of the drug use disorders identification test (DUDIT): a review of recent research. Journal of substance abuse treatment 2015;53:52–59.

33. Bourne A, Reid D, Hickson F, Torres-Rueda S, Weatherburn P. Illicit drug use in sexual settings (‘chemsex’) and HIV/STI transmission risk behaviour among gay men in South London: findings from a qualitative study. Sexually transmitted infections 2015;91(8):564–568.

34. Reinert DF, Allen JP. The alcohol use disorders identification test: an update of research findings. Alcohol Clin Exp Res 2007;31(2):185–99.

35. Kayaert L, Sarink D, Visser M, et al. Sexually transmitted infections in the Netherlands in 2022. Seksueel overdraagbare aandoeningen in Nederland in 2022 Rijksinstituut voor Volksgezondheid en Milieu RIVM, 2023.

36. Solomon MM, Mayer KH, Glidden DV, et al. Syphilis predicts HIV incidence among men and transgender women who have sex with men in a preexposure prophylaxis trial. Clinical Infectious Diseases 2014;59(7):1020–1026.

37. Bernstein KT, Marcus JL, Nieri G, Philip SS, Klausner JD. Rectal gonorrhea and chlamydia reinfection is associated with increased risk of HIV seroconversion. JAIDS Journal of Acquired Immune Deficiency Syndromes 2010;53(4):537–543.

38. Jongen VW, Daans C, van Sighem A, et al. Assessing the HIV care continuum among transgender women during 11 years of follow-up: results from the Netherlands’ ATHENA observational cohort. Journal of the International AIDS Society 2024;27(8):e26317.

39. Stutterheim SE, van Dijk M, Wang H, Jonas KJ. The worldwide burden of HIV in transgender individuals: an updated systematic review and meta-analysis. PloS one 2021;16(12):e0260063.

40. Santoso D, Asfia SK, Mello MB, et al. HIV prevalence ratio of international migrants compared to their native-born counterparts: a systematic review and meta-analysis. EClinicalMedicine 2022;53.

41. Tieosapjaroen W, Zhang Y, Fairley CK, et al. Improving access to oral pre-exposure prophylaxis for HIV among international migrant populations. The Lancet Public Health 2023;8(8):e651–e658.

42. Wirtz AL, Humes E, Althoff KN, et al. HIV incidence and mortality in transgender women in the eastern and southern USA: a multisite cohort study. The Lancet HIV 2023;10(5):e308–e319.

43. Estrela M, Semedo G, Roque F, Ferreira PL, Herdeiro MT. Sociodemographic determinants of digital health literacy: a systematic review and meta-analysis. International Journal of Medical Informatics 2023;177:105124.

44. Plantinga S, Kaal M. Hoe mediawijs is Nederland? Mediawijzer. Kantar Public 2018.

45. Reiners F, Sturm J, Bouw LJ, Wouters EJ. Sociodemographic factors influencing the use of eHealth in people with chronic diseases. International journal of environmental research and public health 2019;16(4):645.

46. Woolley KE, Bright D, Ayres T, et al. Mapping inequities in digital health technology within the World Health Organization’s European Region using PROGRESS PLUS: scoping review. Journal of Medical Internet Research 2023;25:e44181.

47. Korenromp EL, Sabin K, Stover J, et al. New HIV infections among key populations and their partners in 2010 and 2022, by world region: a multisources estimation. JAIDS Journal of Acquired Immune Deficiency Syndromes 2024;95(1S):e34–e45.

48. Beyrer C, Kamarulzaman A, Isbell M, et al. Under threat: the International AIDS Society–Lancet Commission on health and human rights. The Lancet 2024;403(10434):1374–1418.

49. Shannon K, Crago A-L, Baral SD, et al. The global response and unmet actions for HIV and sex workers. The Lancet 2018;392(10148):698–710.

50. Van Mieghem H, Nöstlinger C, Smekens T, et al. Identifying priority groups for pre-exposure prophylaxis among sex workers in Flanders, Belgium: insights into routine HIV and sexually transmitted infection data in community-based clinics. Sexually Transmitted Infections 2024;100(4):236–241.

