## Supplementary Tables for "Representativeness of participants in a randomized controlled trial on modalities of monitoring oral HIV pre-exposure prophylaxis use"

**List of Supplementary Tables:**

**Supplementary Table 1a:** Baseline socio-demographic characteristics and sexual behaviour of EZI-PrEP participants and other PrEP users of the Dutch National PrEP pilot Programme at the Sexual Health Centre in Amsterdam, September 2021 - August 2022

**Supplementary Table 1b:** Baseline socio-demographic characteristics and sexual behaviour of EZI-PrEP participants and other PrEP users of the Dutch National PrEP pilot Programme at the Sexual Health Centre in Rotterdam, September 2021 - August 2022

**Supplementary Table 1c:** Baseline socio-demographic characteristics and sexual behaviour of EZI-PrEP participants and other PrEP users of the Dutch National PrEP pilot Programme at the Sexual Health Centre in the Hague, September 2021 - August 2022

**Supplementary Table 1d:** Baseline socio-demographic characteristics and sexual behaviour of EZI-PrEP participants and other PrEP users of the Dutch National PrEP pilot Programme at the Sexual Health Centre in Nijmegen, September 2021 - August 2022

**Supplementary Table 1: Baseline socio-demographic characteristics, sexual behaviour, STI prevalence and mental health of participants by Sexual Health Centre, EZI-PrEP study, the Netherlands, September 2021 - August 2022**

| | Amsterdam (n=252) | | Rotterdam (n=112) | | The Hague (n=75) | | Nijmegen (n=30) | | |
|---|---|---|---|---|---|---|---|---|---|
| | *n*[1] | *%*[1] | *n*[1] | *%*[1] | *n*[1] | *%*[1] | *n*[1] | *%*[1] | *p-value*[2] |
| **Demographic characteristics** | | | | | | | | | |
| **Gender** | | | | | | | | | |
| Male | 247 | 98% | 112 | 100% | 75 | 100% | 30 | 100% | 0.446 |
| Transgender or gender diverse | 5 | 2% | 0 | 0% | 0 | 0% | 0 | 0% | |
| **Age in years** | | | | | | | | | |
| Median, [IQR] | 37 | [30-47] | 32 | [27-41] | 38 | [32-48] | 37 | [26-53] | <0.001 |
| <25 years | 18 | 7% | 20 | 18% | 4 | 5% | 6 | 20% | 0.004 |
| 25-34 years | 84 | 33% | 48 | 43% | 28 | 37% | 8 | 27% | |
| 35-44 years | 68 | 27% | 22 | 20% | 16 | 21% | 5 | 17% | |
| ≥45 years | 82 | 33% | 22 | 20% | 27 | 36% | 11 | 37% | |
| **Country or region of birth** | | | | | | | | | |
| The Netherlands | 152 | 60% | 83 | 74% | 56 | 75% | 28 | 93% | <0.001[3] |
| Western and Central Europe and North America | 36 | 14% | 9 | 8% | 8 | 11% | 2 | 7% | |

| | | | | | | | | | |
|---|---|---|---|---|---|---|---|---|---|
| Eastern Europe and Central Asia | 14 | 6% | 0 | 0% | 0 | 0% | 0 | 0% | |
| Middle East and North Africa | 10 | 4% | 1 | 1% | 0 | 0% | 0 | 0% | |
| Latin America and the Caribbean | 22 | 9% | 9 | 8% | 8 | 11% | 0 | 0% | |
| Asia and the Pacific | 12 | 5% | 8 | 7% | 2 | 3% | 0 | 0% | |
| Sub-Saharan Africa | 6 | 2% | 2 | 2% | 1 | 1% | 0 | 0% | |
| **Highest completed or current education level[4]** | | | | | | | | | |
| None, primary or other | 5 | 2% | 0 | 0% | 5 | 7% | 2 | 7% | 0.029 |
| Secondary | 37 | 15% | 20 | 18% | 8 | 11% | 7 | 23% | |
| College/university | 205 | 83% | 91 | 82% | 60 | 82% | 21 | 70% | |
| Missing / unknown | 5 | | 1 | | 2 | | 0 | | |
| **Not having a paid job[5]** | 23 | 10% | 13 | 13% | 8 | 11% | 3 | 10% | 0.858 |
| **Income below minimum wage[6]** | 39 | 18% | 22 | 23% | 10 | 16% | 6 | 22% | 0.603 |
| **PrEP use past year** | | | | | | | | | |
| No use | 23 | 9% | 11 | 10% | 14 | 19% | 6 | 20% | 0.076 |
| Used PrEP 4-12 months prior to inclusion | 5 | 2% | 0 | 0% | 2 | 3% | 0 | 0% | |
| Used PrEP in the 3 months before inclusion | 224 | 89% | 101 | 90% | 59 | 79% | 24 | 80% | |
| **Sexual behaviour in preceding 6 months** | | | | | | | | | |
| **Number of sex partners, median [IQR][7]** | 7 | [4-15] | 8 | [4-15] | 6 | [3-10] | 9 | [5-20] | 0.145 |
| **Had insertive anal sex** | 227 | 90% | 93 | 83% | 62 | 83% | 24 | 80% | 0.113 |

| | | | | | | | | | |
|---|---|---|---|---|---|---|---|---|---|
| **Had receptive anal sex** | 205 | 81% | 100 | 89% | 70 | 93% | 25 | 83% | 0.037 |
| **Had condomless anal sex** | 245 | 97% | 103 | 92% | 70 | 93% | 29 | 97% | 0.130 |
| **Had condomless insertive anal sex** | 223 | 88% | 88 | 79% | 59 | 79% | 24 | 80% | 0.042 |
| **Had condomless receptive anal sex** | 202 | 80% | 96 | 86% | 66 | 88% | 24 | 80% | 0.324 |
| **Had group sex**[8] | 66 | 30% | 48 | 43% | 32 | 43% | 22 | 73% | <0.001 |
| **Did sex work**[9] | 0 | 0% | 3 | 3% | 0 | 0% | 0 | 0% | 0.074 |
| **Had chemsex**[10] | 68 | 27% | 27 | 24% | 16 | 21% | 12 | 40% | 0.239 |
| **Injected drugs during or around sex**[11] | 3 | 1% | 0 | 0% | 0 | 0% | 0 | 0% | 0.793 |
| **Bacterial STI diagnoses at inclusion** | | | | | | | | | |
| **Any bacterial STI**[12] | 48 | 20% | 16 | 15% | 14 | 19% | 6 | 20% | 0.696 |
| ***Neisseria gonorrhoeae***[13] | 26 | 11% | 9 | 8% | 6 | 8% | 4 | 13% | 0.732 |
| ***Chlamydia trachomatis***[14] | 22 | 9% | 7 | 6% | 8 | 11% | 3 | 10% | 0.715 |
| **Infectious syphilis**[15] | 6 | 2% | 1 | 1% | 1 | 1% | 0 | 0% | 0.829 |
| **Mental well-being** | | | | | | | | | |
| **Symptoms of depression**[16] | 108 | 46% | 52 | 52% | 28 | 41% | 15 | 52% | 0.521 |
| **Indication for sexual compulsivity**[17] | 45 | 19% | 21 | 20% | 17 | 24% | 5 | 17% | 0.781 |
| **AUDIT categories (score range)** | | | | | | | | | |
| No or low risk alcohol use (0-7) | 158 | 66% | 63 | 61% | 54 | 76% | 20 | 69% | 0.362 |
| Hazardous alcohol use (8-15) | 69 | 29% | 31 | 30% | 15 | 21% | 6 | 21% | |

| | | | | | | | | | |
|---|---|---|---|---|---|---|---|---|---|
| Harmful alcohol use (16-19) | 8 | 3% | 5 | 5% | 1 | 1% | 1 | 3% | |
| Likely dependent on alcohol (≥ 20) | 4 | 2% | 5 | 5% | 1 | 1% | 2 | 7% | |
| Missing | 13 | | 8 | | 4 | | 1 | | |
| **DUDIT categories (score range)** | | | | | | | | | |
| No drugs related problems (0-5) | 130 | 54% | 65 | 64% | 43 | 61% | 15% | 52% | 0.502 |
| Harmful drug use (6-24) | 104 | 44% | 35 | 34% | 28 | 39% | 14% | 48% | |
| Likely dependent on drugs (≥ 25) | 5 | 2% | 2 | 2% | 0 | 0% | 0% | 0% | |
| Missing | 13 | | 10 | | 4 | | 1 | | |

*1. Unless stated otherwise*

*2. Based on Kruskal-Wallis tests for continuous variables and Pearson's $\chi^2$ or Fisher's exact test for categorical variables.*

*3. P-value is based on binary comparison of individuals born in the Netherlands versus not born in the Netherlands.*

*4. None or Primary or other: no education, elementary school, lbo, mavo, vmbo, mbo-1; Secondary: mbo-2-4, havo, vwo; University or College: university of applied sciences, university. Binary comparison of college/university vs. secondary, none/primary/other: Amsterdam 83% (n=205), Rotterdam 82% (n=91), The Hague 82% (n=60), Nijmegen 70% (n=21), p-value 0.384.*

*5. Included in not having a paid job: student/school (n=23), volunteer (n=3), unemployed (n=10), unable to work (n=4), retired (n=7). Missing/unkown: Amsterdam (n=13), Rotterdam (n=10), The Hague (n=4), Nijmegen (n=1).*

*6. Minimum wage 2021: € 1701 gross monthly income. Missing/not willing to disclose: Amsterdam (n=36), Rotterdam (n=18), The Hague (n=12), Nijmegen (n=3).*

*7. Missing/unkown: Amsterdam (n=6), Rotterdam (n=0), The Hague (n=6), Nijmegen (n=0).*

*8. Missing/unkown: Amsterdam (n=30), Rotterdam (n=1), The Hague (n=0), Nijmegen (n=0).*

*9. Missing/unkown: Amsterdam (n=7), Rotterdam (n=0), The Hague (n=14), Nijmegen (n=0).*

*10. Chemsex: sex under influence of one or more of the following drugs: crystal methamphetamine; mephedrone; GHB/GBL; ketamine. Missing/unkown: Amsterdam (n=1), Rotterdam (n=0), The Hague (n=0) , Nijmegen (n=0).*

*11. Drugs included: 3-mmc; heroin; ketamine; cocaine. Missing/unkown: Amsterdam (n=1), Rotterdam (n=0), The Hague (n=0), Nijmegen (n=0).*

*12. Having at least 1 bacterial STI. Missing/unkown: Amsterdam (n=12), Rotterdam (n=3), The Hague (n=0), Nijmegen (n=0).*

*13. Missing/unknown: Amsterdam (n=13), Rotterdam (n=2), The Hague (n=0), Nijmegen (n=0)*

*14. Missing/unknown: Amsterdam (n=13), Rotterdam (n=3), The Hague (n=0), Nijmegen (n=0)*

*15. Infectious syphilis: Lues I, Lues II, Lues latens recens. Missing/unknown: Amsterdam (n=7), Rotterdam (n=2), The Hague (n=0), Nijmegen (n=0).*

*16. Patient Health Questionnaire-9 (PHQ- 9) score ≥5. Missing/unknown: Amsterdam (n=18), Rotterdam (n=12), The Hague (n=7), Nijmegen (n=1).*

*17. Sexual compulsivity scale (SCS) score ≥24. Missing/unknown: Amsterdam (n=11), Rotterdam (n=7), The Hague (n=4), Nijmegen (n=1).*

*Note: Proportions are based on complete data; missing/unknown data excluded from calculations.*

*Abbreviations: EZI-PrEP: E-Health for Zero Infections - facilitating access to and use of Pre-Exposure Prophylaxis in the Netherlands; IQR: interquartile range; PrEP: pre-exposure prophylaxis; STI: sexually transmitted infection; AUDIT: Alcohol Use Disorders Identification Test; DUDIT: Drug Use Disorders Identification Test.*

**Supplementary Table 2: Baseline socio-demographic characteristics and sexual behaviour of EZI-PrEP participants and other PrEP users of the Dutch National PrEP pilot Programme at the Sexual Health Centres in Amsterdam, Rotterdam, The Hague and Nijmegen, September 2021 to August 2022**

| | EZI-PrEP (n=469) | | NPP (n=5161) | | |
|---|---|---|---|---|---|
| | *n*[1] | *%*[1] | *n*[1] | *%*[1] | *p-value*[2] |
| **Sexual Health Center** | | | | | |
| Amsterdam | 252 | 54% | 3,030 | 59% | 0.003 |
| Rotterdam | 112 | 24% | 1,234 | 24% | |
| The Hague | 75 | 16% | 543 | 11% | |
| Nijmegen | 30 | 6% | 360 | 7% | |
| **Demographic characteristics** | | | | | |
| **Gender** | | | | | |
| Male | 464 | 99% | 4954 | 96% | 0.001 |
| Transgender or gender diverse | 5 | 1% | 207 | 4% | |
| **Age in years** | | | | | |
| Median, [IQR] | 36 | [29-47] | 34 | [28-44] | 0.005 |
| <25years[3] | 48 | 10% | 655 | 12% | 0.046 |
| 25-34 years | 168 | 36% | 1,975 | 38% | |
| 35-44 years | 111 | 24% | 1,253 | 24% | |
| ≥45 years | 142 | 30% | 1,278 | 25% | |
| **Country of birth** | | | | | |

| | | | | | |
|---|---|---|---|---|---|
| The Netherlands[4] | 319 | 68% | 2,982 | 58% | <0.001 |
| Western and Central Europe and North America | 55 | 12% | 548 | 11% | |
| Eastern Europe and Central Asia | 14 | 3% | 263 | 5% | |
| Middle East and North Africa | 11 | 2% | 370 | 7% | |
| Latin America and the Caribbean | 39 | 8% | 627 | 12% | |
| Asia and the Pacific | 22 | 5% | 275 | 5% | |
| Sub-Saharan Africa | 9 | 2% | 83 | 2% | |
| Missing / unknown | 1 | | 13 | | |
| **Highest completed or current education level[5]** | | | | | |
| None, primary or other | 12 | 3% | 321 | 6% | 0.001 |
| Secondary | 72 | 16% | 791 | 17% | |
| College/university | 377 | 82% | 3,562 | 77% | |
| Missing / unknown | 8 | | 487 | | |
| **PrEP use past year** | | | | | |
| No use | 54 | 12% | 565 | 11% | 0.402 |
| Used PrEP 4-12 months prior to inclusion | 7 | 1% | 126 | 2% | |
| Used PrEP in the 3 months before inclusion | 408 | 87% | 4,389 | 86% | |
| Unknown/missing | 0 | | 81 | | |
| **Sexual behaviour in preceding 6 months** | | | | | |
| **Number of sex partners, median [IQR][6]** | 7 | [4-15] | 6 | [3-14] | 0.056 |
| **Had insertive anal sex** | 406 | 87% | 4,404 | 85% | 0.468 |
| **Had receptive anal sex** | 400 | 85% | 4,329 | 84% | 0.426 |

| | | | | | |
|---|---|---|---|---|---|
| **Had condomless anal sex** | 447 | 95% | 4,726 | 92% | 0.005 |
| **Had condomless insertive anal sex** | 394 | 84% | 4,092 | 79% | 0.015 |
| **Had condomless receptive anal sex** | 388 | 83% | 4,018 | 88% | 0.014 |
| **Had group sex** [7] | 168 | 38% | 1672 | 33% | 0.024 |
| **Did sex work** [8] | 3 | 1% | 291 | 6% | <0.001 |
| **Had chemsex**[9] | 123 | 26% | 1,157 | 22% | 0.060 |
| **Injected drugs during or around sex**[10] | 3 | 1% | 55 | 1% | 0.276 |
| **Bacterial STI diagnoses** | | | | | |
| **Any bacterial STI**[11] | 84 | 19% | 894 | 18% | 0.766 |
| ***Neisseria gonorrhoea***[12] | 45 | 10% | 524 | 11% | 0.696 |
| ***Chlamydia trachomatis***[13] | 40 | 9% | 443 | 9% | 0.973 |
| **Infectious syphilis**[14] | 8 | 2% | 78 | 2% | 0.737 |

*1. Unless stated otherwise*

*2. Based on Wilcoxon rank-sum tests for continuous variables and Pearson's $\chi^2$ or Fisher's exact for categorical variables.*

*3. Binary comparison of those under 25 year old, p-value 0.123.*

*4. Binary comparison of those born in the Netherlands, p-value < 0.001.*

*5. None or Primary or other: no education, elementary school, lbo, mavo, vmbo, mbo-1; Secondary: mbo-2-4, havo, vwo; University or College: university of applied sciences, university. Binary comparison of college/university vs. secondary,none/primary/other, p-value 0.007.*

*6. Missing/unkown: EZI-PrEP (n=12), NPP (n=90).*

*7. Missing/unkown: EZI-PrEP (n=31), NPP (n=102).*

*8. Missing/unkown: EZI-PrEP (n=21), NPP (n=570).*

*9. Chemsex sex under influence of one or more of the following drugs: crystal methamphetamine; mephedrone; GHB/GBL; ketamine. Missing/unkown: EZI-PrEP (n=1),NPP (n=13).*

*10. Drugs included: 3-mmc; heroin; ketamine; cocaine. Missing/unkown: EZI-PrEP (n=1),NPP (n=13).*

*11. Having at least 1 bacterial STI. Missing/unknown: EZI-PrEP (n=15),NPP (n=178).*

*12. Missing/unknown: EZI-PrEP (n=15),NPP (n=169).*

*13. Missing/unknown: EZI-PrEP (n=16),NPP (n=171).*

*14. Infectious syphilis: Lues I, Lues II, Lues latens recens. Missing/unkown: EZI-PrEP (n=9),NPP (n=87).*

*Note: Proportions are based on complete data; missing/unknown data excluded from calculations.*

*EZI-PrEP: E-Health for Zero Infections - facilitating access to and use of Pre-Exposure Prophylaxis in the Netherlands; IQR: interquartile range; PrEP: pre-exposure prophylaxis; STI: sexually transmitted infection.*

**Supplementary Table 3a: Baseline socio-demographic characteristics and sexual behaviour of EZI-PrEP participants and other PrEP users of the Dutch National PrEP pilot Programme at the Sexual Health Centre in Amsterdam, September 2021 - August 2022**

| | EZI-PrEP Amsterdam (n=252) | | NPP Amsterdam (n=3026) | | |
|---|---|---|---|---|---|
| | *n[1]* | *%[1]* | *n[1]* | *%[1]* | *p-value[2]* |
| **Demographic characteristics** | | | | | |
| **Gender** | | | | | |
| Male | 247 | 98% | 2,861 | 95% | 0.017 |
| Transgender or gender diverse | 5 | 2% | 165 | 5% | |
| **Age in years** | | | | | |
| Median, [IQR] | 37 | [30-47] | 33 | [28-43] | <0.001 |
| <25years[3] | 18 | 7% | 378 | 12% | <0.001 |
| 25-34 years | 84 | 33% | 1,238 | 41% | |
| 35-44 years | 68 | 27% | 733 | 24% | |
| ≥45 years | 82 | 33% | 677 | 22% | |
| **Country of birth** | | | | | |
| The Netherlands[4] | 152 | 60% | 1,534 | 51% | 0.013 |
| Western and Central Europe and North America | 36 | 14% | 410 | 14% | |
| Eastern Europe and Central Asia | 14 | 6% | 175 | 6% | |
| Middle East and North Africa | 10 | 4% | 275 | 9% | |

| | | | | | |
|---|---|---|---|---|---|
| Latin America and the Caribbean | 22 | 9% | 394 | 13% | |
| Asia and the Pacific | 12 | 5% | 181 | 6% | |
| Sub-Saharan Africa | 6 | 2% | 47 | 2% | |
| Missing / unknown | 0 | | 10 | | |
| **Highest completed or current education level[5]** | | | | | |
| None, primary or other | 5 | 2% | 189 | 7% | 0.005 |
| Secondary | 37 | 15% | 321 | 12% | |
| College/university | 205 | 83% | 2,114 | 81% | |
| Missing / unknown | 5 | | 402 | | |
| **PrEP use past year** | | | | | |
| No use | 23 | 9% | 314 | 11% | 0.656 |
| Used PrEP 4-12 months prior to inclusion | 5 | 2% | 74 | 3% | |
| Used PrEP in the 3 months before inclusion | 224 | 89% | 2,572 | 87% | |
| Unknown/missing | 0 | | 66 | | |
| **Sexual behaviour in preceding 6 months** | | | | | |
| **Number of sex partners, median [IQR][6]** | 7 | [4-15] | 6 | [3-15] | 0.164 |
| **Had insertive anal sex** | 227 | 90% | 2,582 | 85% | 0.038 |
| **Had receptive anal sex** | 205 | 81% | 2,479 | 82% | 0.820 |
| **Had condomless anal sex** | 245 | 97% | 2,743 | 91% | <0.001 |
| **Had condomless insertive anal sex** | 223 | 88% | 2,380 | 79% | <0.001 |
| **Had condomless receptive anal sex** | 202 | 80% | 2,292 | 76% | 0.114 |
| **Had group sex[7]** | 66 | 30% | 791 | 27% | 0.335 |
| **Did sex work[8]** | 0 | 0% | 241 | 10% | <0.001 |

| | | | | | |
|---|---|---|---|---|---|
| **Had chemsex[9]** | 68 | 27% | 699 | 23% | 0.160 |
| **Injected drugs during or around sex[10]** | 3 | 1% | 32 | 1% | 0.748 |
| **Bacterial STI diagnoses** | | | | | |
| **Any bacterial STI[11]** | 48 | 20% | 556 | 19% | 0.741 |
| ***Neisseria gonorrhoeae*[12]** | 26 | 11% | 344 | 12% | 0.661 |
| ***Chlamydia trachomatis*[13]** | 22 | 9% | 268 | 9% | 0.994 |
| **Infectious syphilis[14]** | 6 | 2% | 47 | 2% | 0.303 |

*1. Unless stated otherwise*

*2. Based on Wilcoxon rank-sum tests for continuous variables and Pearson's $\chi^2$ or Fisher's exact for categorical variables.*

*3. Binary comparison of those under 25 year old, p-value 0.012.*

*4. Binary comparison of those born in the Netherlands, p-value 0.004.*

*5. None or Primary or other: no education, elementary school, lbo, mavo, vmbo, mbo-1; Secondary: mbo-2-4, havo, vwo; University or College: university of applied sciences, university. Binary comparison of college/university vs. secondary, none/primary/other, p-value 0.354.*

*6. Missing/unkown: EZI-PrEP (n=6), NPP (n=66).*

*7. Missing/unkown: EZI-PrEP (n=30), NPP (n=69).*

*8. Missing/unkown: EZI-PrEP (n=7), NPP (n=545).*

*9. Chemsex: sex under influence of one or more of the following drugs: crystal methamphetamine; mephedrone; GHB/GBL; ketamine. Missing/unkown: EZI-PrEP (n=1),NPP (n=10).*

*10. Drugs included: 3-mmc; heroin; ketamine; cocaine. Missing/unkown: EZI-PrEP (n=1),NPP (n=10).*

*11. Having at least 1 bacterial STI. Missing/unknown: EZI-PrEP (n=12),NPP (n=119).*

*12. Missing/unknown: EZI-PrEP (n=13),NPP (n=118).*

*13. Missing/unknown: EZI-PrEP (n=13),NPP (n=119).*

*14. Infectious syphilis: Lues I, Lues II, Lues latens recens. Missing/unkown: EZI-PrEP (n=7),NPP (n=49).*

*Note: Proportions are based on complete data; missing/unknown data excluded from calculations.*

*EZI-PrEP: E-Health for Zero Infections - facilitating access to and use of Pre-Exposure Prophylaxis in the Netherlands; IQR: interquartile range; PrEP: pre-exposure prophylaxis; STI: sexually transmitted infection.*

**Supplementary Table 3b: Baseline socio-demographic characteristics and sexual behaviour of EZI-PrEP participants and other PrEP users of the Dutch National PrEP pilot Programme at the Sexual Health Centre in Rotterdam, September 2021 - August 2022**

| | **EZI-PrEP Rotterdam (n=112)** | | **NPP Rotterdam (n=1233)** | | |
|---|---|---|---|---|---|
| | *n*[1] | *%*[1] | *n*[1] | *%*[1] | *p-value*[2] |
| **Demographic characteristics** | | | | | |
| **Gender** | | | | | |
| Male | 112 | 100% | 1,216 | 99% | 0.389 |
| Transgender or gender diverse | 0 | 0% | 17 | 1% | |
| **Age in years** | | | | | |
| Median, [IQR] | 32 | 27-41 | 35 | 28-43 | 0.058 |
| <25years[3] | 20 | 18% | 150 | 12% | 0.098 |
| 25-34 years | 48 | 43% | 460 | 37% | |
| 35-44 years | 22 | 20% | 336 | 27% | |
| ≥45 years | 22 | 20% | 287 | 23% | |
| **Country of birth** | | | | | |
| The Netherlands[4] | 83 | 74% | 806 | 65% | 0.011 |
| Western and Central Europe and North America | 9 | 8% | 80 | 6% | |
| Eastern Europe and Central Asia | 0 | 0% | 54 | 4% | |
| Middle East and North Africa | 1 | 1% | 60 | 5% | |

| | | | | | |
|---|---|---|---|---|---|
| Latin America and the Caribbean | 9 | 8% | 159 | 13% | |
| Asia and the Pacific | 8 | 7% | 51 | 4% | |
| Sub-Saharan Africa | 2 | 2% | 22 | 2% | |
| Missing / unknown | 0 | | 1 | | |
| **Highest completed or current education level**[5] | | | | | |
| None, primary or other | 0 | 0% | 49 | 4% | 0.013 |
| Secondary | 20 | 18% | 289 | 24% | |
| College/university | 91 | 82% | 868 | 72% | |
| Missing / unknown | 1 | | 27 | | |
| **PrEP use past year** | | | | | |
| No use | 11 | 10% | 63 | 5% | 0.067 |
| Used PrEP 4-12 months prior to inclusion | 0 | 0% | 18 | 1% | |
| Used PrEP in the 3 months before inclusion | 101 | 90% | 1,149 | 93% | |
| Unknown/missing | 0 | | 3 | | |
| **Sexual behaviour in preceding 6 months** | | | | | |
| **Number of sex partners, median [IQR]**[6] | 8 | [4-15] | 6 | [4-12] | 0.278 |
| **Had insertive anal sex** | 93 | 83% | 1,066 | 86% | 0.315 |
| **Had receptive anal sex** | 100 | 89% | 1,059 | 86% | 0.319 |
| **Had condomless anal sex** | 103 | 92% | 1,158 | 94% | 0.413 |
| **Had condomless insertive anal sex** | 88 | 79% | 1,005 | 82% | 0.446 |
| **Had condomless receptive anal sex** | 96 | 86% | 986 | 80% | 0.142 |
| **Had group sex**[7] | 48 | 43% | 525 | 43% | 0.971 |
| **Did sex work**[8] | 3 | 3% | 24 | 2% | 0.488 |

| | | | | | |
|---|---|---|---|---|---|
| **Had chemsex[9]** | 27 | 24% | 269 | 22% | 0.582 |
| **Injected drugs during or around sex[10]** | 0 | 0% | 8 | 1% | 1.000 |
| **Bacterial STI diagnoses** | | | | | |
| **Any bacterial STI[11]** | 16 | 15% | 194 | 16% | 0.685 |
| ***Neisseria gonorrhoeae*[12]** | 9 | 8% | 108 | 9% | 0.785 |
| ***Chlamydia trachomatis*[13]** | 7 | 6% | 97 | 8% | 0.547 |
| **Infectious syphilis[14]** | 1 | 1% | 15 | 1% | 1.000 |

*1. Unless stated otherwise*

*2. Based on Wilcoxon rank-sum tests for continuous variables and Pearson's χ2 or Fisher's exact for categorical variables.*

*3. Binary comparison of those under 25 year old, p-value 0.083.*

*4. Binary comparison of those born in the Netherlands, p-value 0.063.*

*5. None or Primary or other: no education, elementary school, lbo, mavo, vmbo, mbo-1; Secondary: mbo-2-4, havo, vwo; University or College: university of applied sciences, university. Binary comparison of college/university vs. secondary, none/primary/other, p-value 0.023.*

*6. Missing/unkown: EZI-PrEP (n=0), NPP (n=1).*

*7. Missing/unkown: EZI-PrEP (n=1), NPP (n=24).*

*8. Missing/unkown: EZI-PrEP (n=0), NPP (n=3).*

*9. Chemsex: sex under influence of one or more of the following drugs: crystal methamphetamine; mephedrone; GHB/GBL; ketamine. Missing/unkown: EZI-PrEP (n=0),NPP (n=2).*

*10. Drugs included: 3-mmc; heroin; ketamine; cocaine. Missing/unkown: EZI-PrEP (n=0),NPP (n=2).*

*11. Having at least 1 bacterial STI. Missing/unknown: EZI-PrEP (n=3),NPP (n=33).*

*12. Missing/unknown: EZI-PrEP (n=2),NPP (n=27).*

*13. Missing/unknown: EZI-PrEP (n=3),NPP (n=28).*

*14. Infectious syphilis: Lues I, Lues II, Lues latens recens. Missing/unkown: EZI-PrEP (n=2),NPP (n=16).*

*Note: Proportions are based on complete data; missing/unknown data excluded from calculations.*

*EZI-PrEP: E-Health for Zero Infections - facilitating access to and use of Pre-Exposure Prophylaxis in the Netherlands; IQR: interquartile range; PrEP: pre-exposure prophylaxis; STI: sexually transmitted infection.*

**Supplementary Table 3c: Baseline socio-demographic characteristics and sexual behaviour of EZI-PrEP participants and other PrEP users of the Dutch National PrEP pilot Programme at the Sexual Health Centre in the Hague, September 2021 - August 2022**

| | **EZI-PrEP The Hague (n=75)** | | **NPP The Hague (n=542)** | | |
|---|---|---|---|---|---|
| | *n*[1] | *%*[1] | *n*[1] | *%*[1] | *p-value*[2] |
| **Demographic characteristics** | | | | | |
| **Gender** | | | | | |
| Male | 75 | 100% | 520 | 96% | 0.095 |
| Transgender or gender diverse | 0 | 0% | 22 | 4% | |
| **Age in years** | | | | | |
| Median, [IQR] | 38 | [32-48] | 34 | [27-46] | 0.012 |
| <25years[3] | 4 | 5% | 87 | 16% | 0.079 |
| 25-34 years | 28 | 37% | 199 | 37% | |
| 35-44 years | 16 | 21% | 105 | 19% | |
| ≥45 years | 27 | 36% | 151 | 28% | |
| **Country of birth** | | | | | |
| The Netherlands[4] | 56 | 75% | 328 | 61% | 0.050 |
| Western and Central Europe and North America | 8 | 11% | 49 | 9% | |
| Eastern Europe and Central Asia | 0 | 0% | 24 | 4% | |
| Middle East and North Africa | 0 | 0% | 27 | 5% | |

| | | | | | |
|---|---|---|---|---|---|
| Latin America and the Caribbean | 8 | 11% | 64 | 12% | |
| Asia and the Pacific | 2 | 3% | 39 | 7% | |
| Sub-Saharan Africa | 1 | 1% | 9 | 2% | |
| Missing / unknown | 0 | | 2 | | |
| **Highest completed or current education level**[5] | | | | | |
| None, primary or other | 5 | 7% | 48 | 10% | 0.526 |
| Secondary | 8 | 11% | 70 | 14% | |
| College/university | 60 | 82% | 379 | 76% | |
| Missing / unknown | 2 | | 45 | | |
| **PrEP use past year** | | | | | |
| No use | 14 | 19% | 163 | 31% | 0.075 |
| Used PrEP 4-12 months prior to inclusion | 2 | 3% | 16 | 3% | |
| Used PrEP in the 3 months before inclusion | 59 | 79% | 353 | 66% | |
| Unknown/missing | 0 | | 10 | | |
| **Sexual behaviour in preceding 6 months** | | | | | |
| **Number of sex partners, median [IQR]**[6] | 6 | [3-10] | 6 | [4-12] | 0.483 |
| **Had insertive anal sex** | 62 | 83% | 451 | 83% | 0.906 |
| **Had receptive anal sex** | 70 | 93% | 486 | 90% | 0.319 |
| **Had condomless anal sex** | 70 | 93% | 497 | 92% | 0.627 |
| **Had condomless insertive anal sex** | 59 | 79% | 421 | 78% | 0.846 |
| **Had condomless receptive anal sex** | 66 | 88% | 452 | 83% | 0.308 |
| **Had group sex**[7] | 32 | 43% | 179 | 33% | 0.109 |

| | | | | | |
|---|---|---|---|---|---|
| **Did sex work[8]** | 0 | 0% | 22 | 4% | 0.153 |
| **Had chemsex[9]** | 16 | 21% | 81 | 15% | 0.154 |
| **Injected drugs during or around sex[10]** | 0 | 0% | 7 | 1% | 1.000 |
| **Bacterial STI diagnoses** | | | | | |
| **Any bacterial STI[11]** | 14 | 19% | 84 | 16% | 0.583 |
| ***Neisseria gonorrhoeae*[12]** | 6 | 8% | 46 | 9% | 0.808 |
| ***Chlamydia trachomatis*[13]** | 8 | 11% | 48 | 9% | 0.691 |
| **Infectious syphilis[14]** | 1 | 1% | 7 | 1% | 1.000 |

*1. Unless stated otherwise*

*2. Based on Wilcoxon rank-sum tests for continuous variables and Pearson's χ2 or Fisher's exact for categorical variables.*

*3. Binary comparison of those under 25 year old, p-value 0.014.*

*4. Binary comparison of those born in the Netherlands, p-value 0.020.*

*5. None or Primary or other: no education, elementary school, lbo, mavo, vmbo, mbo-1; Secondary: mbo-2-4, havo, vwo; University or College: university of applied sciences, university. Binary comparison of college/university vs. secondary, none/primary/other, p-value 0.260.*

*6. Missing/unkown: EZI-PrEP (n=6), NPP (n=26).*

*7. Missing/unkown: EZI-PrEP (n=0), NPP (n=4).*

*8. Missing/unkown: EZI-PrEP (n=14), NPP (n=26).*

*9. Chemsex: sex under influence of one or more of the following drugs: crystal methamphetamine; mephedrone; GHB/GBL; ketamine. Missing/unkown: EZI-PrEP (n=0),NPP (n=0).*

*10. Drugs included: 3-mmc; heroin; ketamine; cocaine. Missing/unkown: EZI-PrEP (n=0),NPP (n=0).*

*11. Having at least 1 bacterial STI. Missing/unknown: EZI-PrEP (n=0),NPP (n=22).*

*12. Missing/unknown: EZI-PrEP (n=0),NPP (n=22).*

*13. Missing/unknown: EZI-PrEP (n=0),NPP (n=22).*

*14. Infectious syphilis: Lues I, Lues II, Lues latens recens. Missing/unkown: EZI-PrEP (n=0),NPP (n=19).*

*Note: Proportions are based on complete data; missing/unknown data excluded from calculations.*

*EZI-PrEP: E-Health for Zero Infections - facilitating access to and use of Pre-Exposure Prophylaxis in the Netherlands; IQR: interquartile range; PrEP: pre-exposure prophylaxis; STI: sexually transmitted infection.*

**Supplementary Table 3d: Baseline socio-demographic characteristics and sexual behaviour of EZI-PrEP participants and other PrEP users of the Dutch National PrEP pilot Programme at the Sexual Health Centre in Nijmegen, September 2021 - August 2022**

| | **EZI-PrEP Nijmegen (n=30)** | | **NPP Nijmegen (n=360)** | | |
|---|---|---|---|---|---|
| | *n*[1] | *%*[1] | *n*[1] | *%*[1] | *p-value*[2] |
| **Demographic characteristics** | | | | | |
| **Gender** | | | | | |
| Male | 30 | 100% | 357 | 99% | 0.616 |
| Transgender or gender diverse | 0 | 0% | 3 | 1% | |
| **Age in years** | | | | | |
| Median, [IQR] | 37 | [26-53] | 43 | [31-53] | 0.169 |
| <25years[3] | 6 | 20% | 40 | 11% | 0.394 |
| 25-34 years | 8 | 27% | 78 | 22% | |
| 35-44 years | 5 | 17% | 79 | 22% | |
| ≥45 years | 11 | 37% | 163 | 45% | |
| **Country of birth** | | | | | |
| The Netherlands[4] | 28 | 93% | 314 | 87% | 0.782 |
| Western and Central Europe and North America | 2 | 7% | 9 | 3% | |
| Eastern Europe and Central Asia | 0 | 0% | 10 | 3% | |
| Middle East and North Africa | 0 | 0% | 8 | 2% | |

| | | | | | |
|---|---|---|---|---|---|
| Latin America and the Caribbean | 0 | 0% | 10 | 3% | |
| Asia and the Pacific | 0 | 0% | 4 | 1% | |
| Sub-Saharan Africa | 0 | 0% | 5 | 1% | |
| Missing / unknown | 0 | | 0 | | |
| **Highest completed or current education level[5]** | | | | | |
| None, primary or other | 2 | 7% | 35 | 10% | 0.510 |
| Secondary | 7 | 23% | 111 | 32% | |
| College/university | 21 | 70% | 201 | 58% | |
| Missing / unknown | 0 | | 13 | | |
| **PrEP use past year** | | | | | |
| No use | 6 | 20% | 25 | 7% | 0.023 |
| Used PrEP 4-12 months prior to inclusion | 0 | 0% | 18 | 5% | |
| Used PrEP in the 3 months before inclusion | 24 | 80% | 315 | 88% | |
| Unknown/missing | 0 | | 2 | | |
| **Sexual behaviour in preceding 6 months** | | | | | |
| **Number of sex partners, median [IQR][6]** | 9 | [5-20] | 5 | [3-10] | 0.013 |
| **Had insertive anal sex** | 24 | 80% | 305 | 85% | 0.494 |
| **Had receptive anal sex** | 25 | 83% | 305 | 85% | 0.839 |
| **Had condomless anal sex** | 29 | 97% | 328 | 91% | 0.294 |
| **Had condomless insertive anal sex** | 24 | 80% | 286 | 79% | 0.942 |
| **Had condomless receptive anal sex** | 24 | 80% | 288 | 80% | 1.000 |
| **Had group sex[7]** | 22 | 73% | 177 | 50% | 0.013 |
| **Did sex work[8]** | 0 | 0% | 4 | 1% | 0.725 |

| | | | | | |
|---|---|---|---|---|---|
| **Had chemsex**[9] | 12 | 40% | 108 | 30% | 0.259 |
| **Injected drugs during or around sex**[10] | 0 | 0% | 8 | 2% | 0.523 |
| **Bacterial STI diagnoses** | | | | | |
| **Any bacterial STI**[11] | 6 | 20% | 60 | 17% | 0.660 |
| ***Neisseria gonorrhoeae***[12] | 4 | 13% | 26 | 7% | 0.273 |
| ***Chlamydia trachomatis***[13] | 3 | 10% | 30 | 8% | 0.732 |
| **Infectious syphilis**[14] | 0 | 0% | 9 | 3% | 1.000 |

*1. Unless stated otherwise*

*2. Based on Wilcoxon rank-sum tests for continuous variables and Pearson's χ2 or Fisher's exact for categorical variables.*

*3. Binary comparison of those under 25 year old, p-value 0.147.*

*4. Binary comparison of those born in the Netherlands, p-value 0.328.*

*5. None or Primary or other: no education, elementary school, lbo, mavo, vmbo, mbo-1; Secondary: mbo-2-4, havo, vwo; University or College: university of applied sciences, university. Binary comparison of college/university vs. secondary, none/primary/other, p-value 0.197.*

*6. Missing/unkown: EZI-PrEP (n=0), NPP (n=3).*

*7. Missing/unkown: EZI-PrEP (n=0), NPP (n=5).*

*8. Missing/unkown: EZI-PrEP (n=0), NPP (n=0).*

*9. Chemsex: sex under influence of one or more of the following drugs: crystal methamphetamine; mephedrone; GHB/GBL; ketamine. Missing/unkown: EZI-PrEP (n=0),NPP (n=1).*

*10. Drugs included: 3-mmc; heroin; ketamine; cocaine. Missing/unkown: EZI-PrEP (n=0),NPP (n=1).*

*11. Having at least 1 bacterial STI. Missing/unknown: EZI-PrEP (n=0),NPP (n=4).*

*12. Missing/unknown: EZI-PrEP (n=0),NPP (n=2).*

*13. Missing/unknown: EZI-PrEP (n=0),NPP (n=2).*

*14. Infectious syphilis: Lues I, Lues II, Lues latens recens. Missing/unkown: EZI-PrEP (n=0),NPP (n=3).*

*Note: Proportions are based on complete data; missing/unknown data excluded from calculations.*

*EZI-PrEP: E-Health for Zero Infections - facilitating access to and use of Pre-Exposure Prophylaxis in the Netherlands; IQR: interquartile range; PrEP: pre-exposure prophylaxis; STI: sexually transmitted infection.*
